# Causal Mediation Pathways in Continuous Postprandial Glucose Monitoring for Type 1 Diabetes Patients

**DOI:** 10.64898/2026.03.16.26348520

**Authors:** Spencer Hilligoss, Annie Qu

## Abstract

Managing postprandial glucose in Type 1 Diabetes Mellitus (T1DM) requires understanding how carbohydrate intake affects glucose through both direct pathways and insulin-mediated compensation. Standard analyses treat insulin as a confounder rather than a mediator, obscuring these distinct causal channels and patient-level heterogeneity in carbohydrate response. Using meal-centered continuous glucose monitoring windows from twelve adults in the OhioT1DM cohorts, we apply causal mediation analysis to decompose the total effect of carbohydrate intake on glucose change into average direct (ADE), insulin-mediated (ACME), and total effects, by meal type and across outcome quantiles. To adjust for pre-meal confounding, we introduce a Causally-constrained Linear Autoencoder that learns low-dimensional representations improving adjustment for measured pre-meal state. Across cohorts, carbohydrate intake raises postprandial glucose chiefly through a large direct effect that insulin only partially offsets. The prespecified primary endpoint, the pooled ADE of a +30 g contrast at two hours, is large and highly significant, robust to multiple-testing correction, and replicates in the independent DiaTrend cohort (54 adults); the insulin-mediated effect is comparatively small. Meal- and quantile-specific signals, including greater dinner-time excursions, do not survive false-discovery-rate correction and are reported as hypothesis-generating. These results localize where postprandial insulin dosing may be refined.

## INTRODUCTION

Type 1 Diabetes Mellitus (T1DM) is an autoimmune condition in which pancreatic *β*-cell destruction leads to absolute insulin deficiency.^1^ Day-to-day management relies on coordinating exogenous insulin with carbohydrate intake to keep glucose within safe limits ^2,3^ and to avoid acute complications such as hypoglycemia ^4–7^ and hyperglycemia, as well as longer-term risks including neuropathy and cardiovascular disease. ^8–13^ In practice, achieving stable postprandial control is difficult as patients and clinicians must align the timing of insulin action with the kinetics of glucose appearance after a meal, ^14,15^ yet these processes often fail to peak together, producing early spikes or late dips. ^16,17^

Multiple sources of variability compound this challenge. Insulin absorption and action exhibit substantial intra- and inter-individual variability.^18^ Additionally, carbohydrate counting and meal composition vary from one event to the next, ^19,20^ while physiological state (sleep, activity, circadian timing) shifts across the day.^21^ Consequently, postprandial glucose responses display clear heterogeneity both between subjects and within the same subject across meals and times of day.^14,16,22,23^ Treatment sizes (both carbohydrate intake and bolus insulin dose) also vary widely, further complicating comparisons and clinical decisionmaking. ^24,25^ A core inferential difficulty is that bolus insulin is typically administered in response to meal carbohydrates and thus lies on the causal pathway from carbohydrates to postprandial glucose. ^26^ Figure 1 is shown for illustration. Disentangling these pathways from observational data is essential for identifying when insulin dosing is insufficient (large direct effects) versus when insulin is effective (substantial mediated effects). ^27^ Glucose-forecasting models and automated insulin-delivery controllers estimate what glucose will do and how much insulin to deliver, but they do not decompose why a postprandial excursion persists. ^28^ Separating the direct effect of carbohydrates from the insulin-mediated effect distinguishes insufficient dosing (a large direct effect) from effective-but-incomplete insulin action (a large mediated effect), a distinction that maps onto different management levers (the insulin-to-carbohydrate ratio versus bolus timing and absorption) and is not recoverable from prediction accuracy alone.

**Figure 1:**
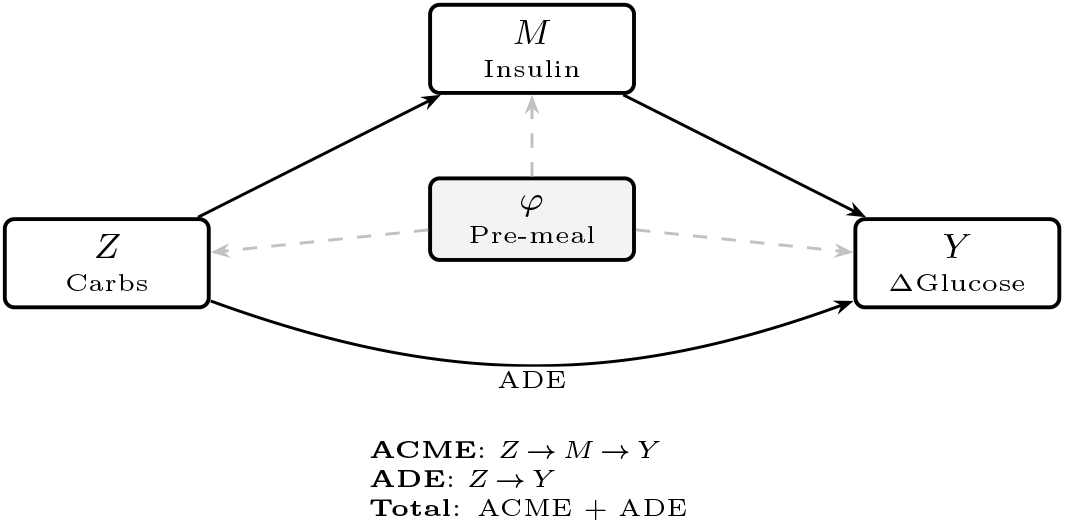
Causal mediation structure for postprandial glucose analysis. The treatment *Z* (meal carbohydrates in grams) affects the outcome *Y* (postprandial glucose change Δ*G* over [+60, +210] minutes) through two pathways: an indirect effect mediated by bolus insulin *M* (aggregated over [ −120, +60] minutes) and a direct effect. Pre-treatment covariates φ, learned from the 2-hour premeal CGM trajectory, adjust for confounding of all three relationships (dashed arrows). The average causal mediation effect (ACME) quantifies the *Z* →*M* → *Y* pathway, while the average direct effect (ADE) captures carbohydrate effects not operating through insulin.

We address this problem using causal mediation analysis (CMA) in the counterfactual framework of Imai et al. ^29,30^. Our analysis focuses on the postprandial change in glucose relative to the pre-meal baseline over a 3.5-hour post-meal horizon and decomposes effects into the average causal mediation effect (ACME), average direct effect (ADE), and total effect (ATE). ^27,31,32^ To capture clinically relevant tail behavior, we model outcomes via quantile regression, allowing effects to vary over time and by quantile (e.g., hypoglycemia-relevant lower tail). We further characterize heterogeneity by meal type and treatment size using clinically interpretable carbohydrate contrasts anchored at the meal-type median. To improve adjustment for individual histories without hand-crafted features, we incorporate pretreatment embeddings learned by a two-stage autoencoder trained on postprandial trajectories. ^33,34^

This design targets the practical questions faced by patients and practitioners: When (over the postprandial window), for whom (across subjects and meals), and at which amount do carbohydrates exert direct effects which dominate indirect mediated effects? Our results reveal time-varying, quantile-specific patterns consistent with clinical experience, e.g., near-complete cancellation of opposing direct and mediated effects at breakfast, contrasted with persistent under-compensation by insulin at dinner, and highlight windows where carbohydrate intake strategies may be refined to better align insulin action with glucose excursions. ^35,36^

## RESULTS

### Model selection and representation quality

We evaluated 60 autoencoder configurations spanning two encoder architectures (CNN, LSTM), five optimizers, and six penalty regimes, each trained with three random seeds (Supplementary Table 13). CNN architectures consistently outperformed LSTM on both outcome prediction and covariate balance. The representation used for all reported analyses is a CNN encoder trained with AdamW and the linearization and balancing penalties.

We distinguish two notions of balance, which were conflated in an earlier version of this manuscript. The first is the embedding’s intrinsic balance score, the degree to which treatment is predictable from the embedding alone (one minus twice the absolute deviation of the treatment-classification AUC from 0.5). The second is the npCBPS-weighted covariate balance achieved in the final mediation sample, the quantity directly relevant to identification. In a factorial ablation ^37^ of the penalty components (Supplementary Table 14), the marginal effects of individual penalties on the intrinsic balance score were small or, for the balancing penalty, negative; the configuration that ranks highest by intrinsic balance score (linearization only) is therefore not the configuration used here. We retained the balancing penalty for its role in treatment balancing and selected the final configuration for the weighted covariate balance it achieves in the mediation sample (0.961, reported below), rather than for the intrinsic balance score (Supplementary Table 15). We now present the ablation as an exploratory comparison of representations.

PCA applied to the resulting 8-dimensional embeddings yielded three orthogonal components capturing approximately 90% of the latent variance, which served as covariates in all downstream models. npCBPS weighting on these components achieved adequate covariate balance, reducing mean absolute treatment correlations by 97.2% (from 0.082 to 0.002) with an effective sample size of 190 of 199 observations (95.5%), indicating that balance was achieved without reliance on extreme weights (Supplementary Table 2). The most imbalanced covariate prior to weighting (PC_3_, *r* = −0.121) was reduced to *r <* 0.001 after weighting, and the weight distribution was well-behaved (median = 0.971, IQR = [0.91, 1.04], 1.5% exceeding 2.0), corresponding to a weighted covariate balance of 0.961 in the mediation sample. These diagnostics confirm that, after weighting, the pipeline achieves the covariate balance required for the downstream causal mediation analysis; we emphasize that this concerns balance on measured premeal covariates and does not establish the absence of unmeasured confounding.

### Pooled mediation effects

We first present the causal mediation analysis pooled across all meal types (*N* = 190), using carbohydrate offsets of +15, +30, and +45 g from the meal-type-specific median as treatment contrasts (Supplementary Figure 4).

The temporal evolution of mediation effects across the postprandial window reveals a consistent pattern. The average direct effect (ADE) of a +30 g carbohydrate increase emerges at 60 minutes, rises to its peak at 120 minutes (ADE = 14.81 mg/dL, 95% CI: 5.55 to 23.65, *p <* 0.001), and remains elevated through 210 minutes. The average causal mediation effect (ACME) through insulin follows a parallel but delayed trajectory: it is non-significant at 60 minutes, becomes significant by 90 minutes, and peaks at 120 minutes (ACME = −9.64 mg/dL, 95% CI: −17.17 to −2.67, *p* = 0.01). This lag in the mediation pathway is consistent with the pharmacokinetics of rapid-acting insulin analogs, which require 60–90 minutes to reach peak action. ^15^ The resulting total effect is positive but modest and generally does not reach statistical significance (total at 120 min = 5.17 mg/dL, 95% CI: −1.41 to 11.69, *p* = 0.13), reflecting the partial but incomplete cancellation of the direct glycemic impact by the compensatory insulin response. The average direct effect of a +30 g contrast at 120 minutes is our prespecified primary endpoint (Methods); it is large and remains significant after BenjaminiHochberg correction under every multiplicity family we considered (within-meal-stratum, pooled, and the full effect grid; Supplementary Material). All remaining effects, including the indirect and total effects and the meal- and quantile-stratified estimates reported below, are secondary and exploratory, and we flag explicitly where their nominal significance does not survive correction.

The dose-response pattern across the +15/+30/+45 g contrasts is approximately linear (Table 3), with both ADE and ACME scaling proportionally to the treatment offset. At the +15 g contrast, the ADE at 120 minutes is 7.35 mg/dL (*p* = 0.002) and the ACME is −4.81 mg/dL (*p* = 0.01). At the +45 g contrast, these effects increase to 21.94 mg/dL (*p* = 0.004) and −14.38 mg/dL (*p* = 0.008), respectively. The proportional scaling of both pathways across dose levels supports the linearity of the underlying mediation structure and suggests that the insulin-to-carbohydrate ratio is approximately constant across the observed range of carbohydrate intake. This pattern of opposing direct and mediated effects, with partial but incomplete insulin compensation, characterizes the fundamental mediation structure across the postprandial window.

The mean-level results presented above characterize the average mediation structure, but the partial cancellation of opposing ADE and ACME pathways raises the question of whether this cancellation is uniform across the conditional glucose response distribution or whether it conceals subgroups for whom the net carbohydrate effect is substantially larger. We address this question using quantile regression at *τ* ∈ { 0.25, 0.50, 0.75 } as the outcome model, replacing the LMER specification while retaining the same Tobit mediator model and npCBPS weights.

At *τ* = 0.25, both the ADE and ACME are diminished relative to the mean-level estimates and are generally nonsignificant across the postprandial window (Table 4). The total effect at 120 minutes is 3.61 mg/dL (95% CI: −6.89 to 13.65, *p* = 0.52), confirming that individuals in the lower quartile of the conditional glucose response distribution experience minimal net glycemic impact from additional carbohydrate intake. At *τ* = 0.50, the mediation structure is broadly consistent with the LMER estimates: the ADE at 120 minutes is 19.71 mg/dL (*p <* 0.001) and the ACME is −10.06 mg/dL (*p <* 0.001), with a significant total effect of 9.66 mg/dL (*p* = 0.04) that already exceeds the nonsignificant LMER estimate at the same timepoint.

The most informative departure from the mean-level analysis occurs at *τ* = 0.75. Here, the ADE is amplified relative to the mean (19.73 mg/dL at 120 minutes, *p* = 0.02), while the ACME is attenuated ( −5.79 mg/dL, *p* = 0.31). The widening gap between direct and mediated effects at this quantile produces a total effect of 13.95 mg/dL (*p* = 0.006), nearly three times the non-significant LMER total effect of 5.17 mg/dL (*p* = 0.13) at the same timepoint. This discrepancy arises because the mean averages over a distribution in which near-zero effects at lower quantiles dilute the clinically meaningful net glucose increases experienced by individuals in the upper quartile of the glucose response distribution. Depending on the starting glucose level, the *τ* = 0.75 total effect of 13.95 mg/dL can exceed the threshold commonly used as a clinically meaningful glucose excursion, ^2^ suggesting that the mean-level analysis may understate the net effect in the upper tail of the response distribution. We caution that this quantile-specific total effect is exploratory: its nominal significance (*p* = 0.006) does not survive Benjamini-Hochberg correction across the exploratory grid, and it is reported as hypothesis-generating rather than as a confirmed subgroup effect.

### Meal-type-stratified mediation analysis

The pooled mediation effects treat the carbohydrate-insulin-glucose pathway as homogeneous across the daily meal cycle. Stratified analyses reveal that this assumption is strongly violated as the mediation structure differs qualitatively across meal types, with breakfast and dinner exhibiting the most pronounced and clinically interpretable patterns. Lunch and snack show negligible mediation structure and are reported in full in the Supplementary Material.

Dinner (*N* = 42) shows the largest sustained mediation effects of any meal type, with both the ACME and ADE exceeding the pooled estimates in absolute magnitude (Supplementary Figure 6). For a +30 g carbohydrate increase, the ADE rises from 15.96 mg/dL at 60 minutes to 35.73 mg/dL at 120 minutes (*p* = 0.006) and remains above 31 mg/dL through 210 minutes. The ACME is consistently negative and significant across the full postprandial horizon, ranging from −12.09 mg/dL at 60 minutes to −22.56 mg/dL at 120 minutes (*p* = 0.01) and remaining near −22 mg/dL at 210 minutes (*p* = 0.01). Critically, unlike the near-cancellation observed at breakfast, the ADE at dinner substantially exceeds the ACME in magnitude, producing positive total effects of 10–14 mg/dL across the 90–210 minute window, though these do not reach significance (*p* = 0.14–0.30) given the smaller sample size. The sustained and large ADE at dinner suggests that evening carbohydrate metabolism produces a prolonged glycemic impact that insulin bolusing does not fully counteract.

The quantile regression analysis at dinner reveals a mediation structure that is remarkably consistent across the conditional glucose distribution, in contrast to the pooled analysis, where effects concentrate at the upper quantiles. At *τ* = 0.25, both the ADE and ACME are large in absolute magnitude (ADE = 38.28 mg/dL, *p* = 0.004; ACME = −27.49 mg/dL, *p <* 0.001), but their near-cancellation produces a non-significant total effect of 10.79 mg/dL (*p* = 0.35), indicating that individuals in the lower tail of the dinner glucose distribution still experience substantial opposing direct and mediated effects. At *τ* = 0.50, the point estimates remain large (ADE = 37.44 mg/dL, *p* = 0.06; ACME = −17.49 mg/dL, *p* = 0.18), though neither reaches significance at the 120-minute horizon. This likely reflects the wider confidence intervals associated with the smaller dinner subsample. The total effect at this quantile is 19.95 mg/dL (*p* = 0.17). At *τ* = 0.75, the total effect reaches 22.03 mg/dL at 120 minutes (*p* = 0.04), with an ADE of 49.46 mg/dL (*p* = 0.008) and ACME of −27.43 mg/dL (*p* = 0.04). Taken at face value, this would indicate that individuals in the upper tail of the dinner glucose response distribution experience net postprandial increases exceeding 20 mg/dL for a +30 g carbohydrate contrast. However, this dinner *τ* = 0.75 total effect (*p* = 0.04, *N* = 42) is exploratory and does not survive Benjamini-Hochberg correction; we therefore present the incomplete-compensation pattern at dinner as a hypothesis-generating signal rather than a confirmed subgroup effect (Figure 3). What is robust, and what the corrected analysis emphasizes, is the large direct effect of carbohydrate intake, which dominates the insulin-mediated effect at dinner.

Breakfast (*N* = 51) provides a revealing contrast to dinner (Supplementary Figure 8). For a +30 g carbohydrate increase, the ADE peaks at 90 minutes (23.46 mg/dL, *p* = 0.01) and the ACME peaks at 120 minutes ( −21.15 mg/dL, *p* = 0.004). Both the direct and indirect effects are individually significant from 60 through 120 minutes (*p <* 0.05); the ACME remains significant through 180 minutes while the ADE attenuates after its 90-minute peak, and by 210 minutes both effects are non-significant (*p >* 0.10). Despite this, their near-cancellation produces total effects that are uniformly non-significant across the full horizon (all *p >* 0.35). The magnitude of both effects at their peaks, roughly double the pooled estimates, indicates that the carbohydrate-glucose and carbohydrateinsulin-glucose pathways are both amplified at breakfast relative to other meal types, yet remain sufficiently well-matched to produce net effects indistinguishable from zero. This pattern is consistent with known diurnal variation in insulin sensitivity,^38^ though the present analysis cannot determine whether the amplification reflects physiological factors such as morning insulin resistance or behavioral factors such as more consistent bolusing practices at breakfast.

The quantile regression analysis at breakfast reinforces this interpretation and provides the sharpest contrast with dinner. At *τ* = 0.25, the ADE is 17.53 mg/dL (*p* = 0.24) and the ACME is −10.28 mg/dL (*p* = 0.38); the total effect is 7.25 mg/dL (*p* = 0.23), indicating that individuals with lower glucose responses experience reduced effects through both pathways. At *τ* = 0.50, the mediation structure mirrors the LMER estimates, with an ADE of 16.40 mg/dL (*p* = 0.03) and an ACME of −17.21 mg/dL (*p* = 0.02), maintaining the near-cancellation pattern with a total effect of −0.81 mg/dL (*p* = 0.88). The most clinically informative finding at breakfast concerns the upper quantile: at *τ* = 0.75, the ACME is −24.58 mg/dL (*p <* 0.001) and the ADE is 23.58 mg/dL (*p* = 0.09), with a total effect of −1.00 mg/dL (*p* = 0.96). The near-cancellation pattern persists even at this quantile, confirming that breakfast bolusing is well-calibrated across the full glucose response distribution. This stands in direct contrast to dinner, where the gap between ADE and ACME widens at *τ* = 0.75 (Figure 3).

Neither lunch (N=58) nor snack (N=39) exhibits a detectable mediation structure at any treatment contrast or quantile level (Supplementary Figures 10–13 and Supplementary Tables 9–12). We discuss possible explanations for these null findings, including greater heterogeneity in snack bolusing behavior and more moderate insulin-to-carbohydrate coupling at midday, in the Discussion.

### Cross-meal comparison and distributional heterogeneity

Taken together, these results demonstrate that the mediation structure is not constant across the day, confirming the heterogeneity observed in the descriptive data (Table 1, Figure 2). The most notable contrast is between dinner and breakfast. At dinner, the direct effect is large and sustained, while the compensatory insulin response, though significant, is insufficient to offset it, producing the largest residual glucose impact of any meal type. At breakfast, the direct glycemic effect of additional carbohydrates is matched by a compensatory insulin response of nearly equal magnitude, yielding total effects close to zero.

**Table 1:**
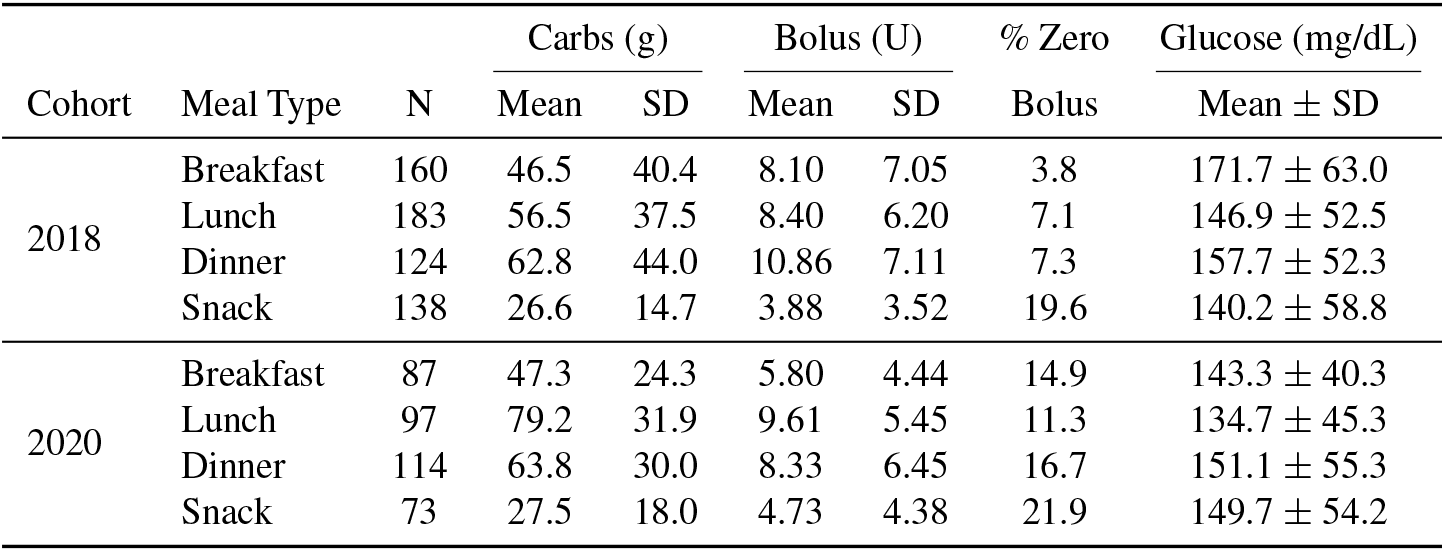
Data characteristics by cohort and meal type. Summary statistics for carbohydrate intake (treatment), insulin bolus (mediator), and pre-meal glucose stratified by data collection cohort and meal category. N = number of meal observations used in the analysis; SD = standard deviation; values are mean *±* SD; % Zero Bolus = percentage of meals with no insulin bolus.

**Figure 2:**
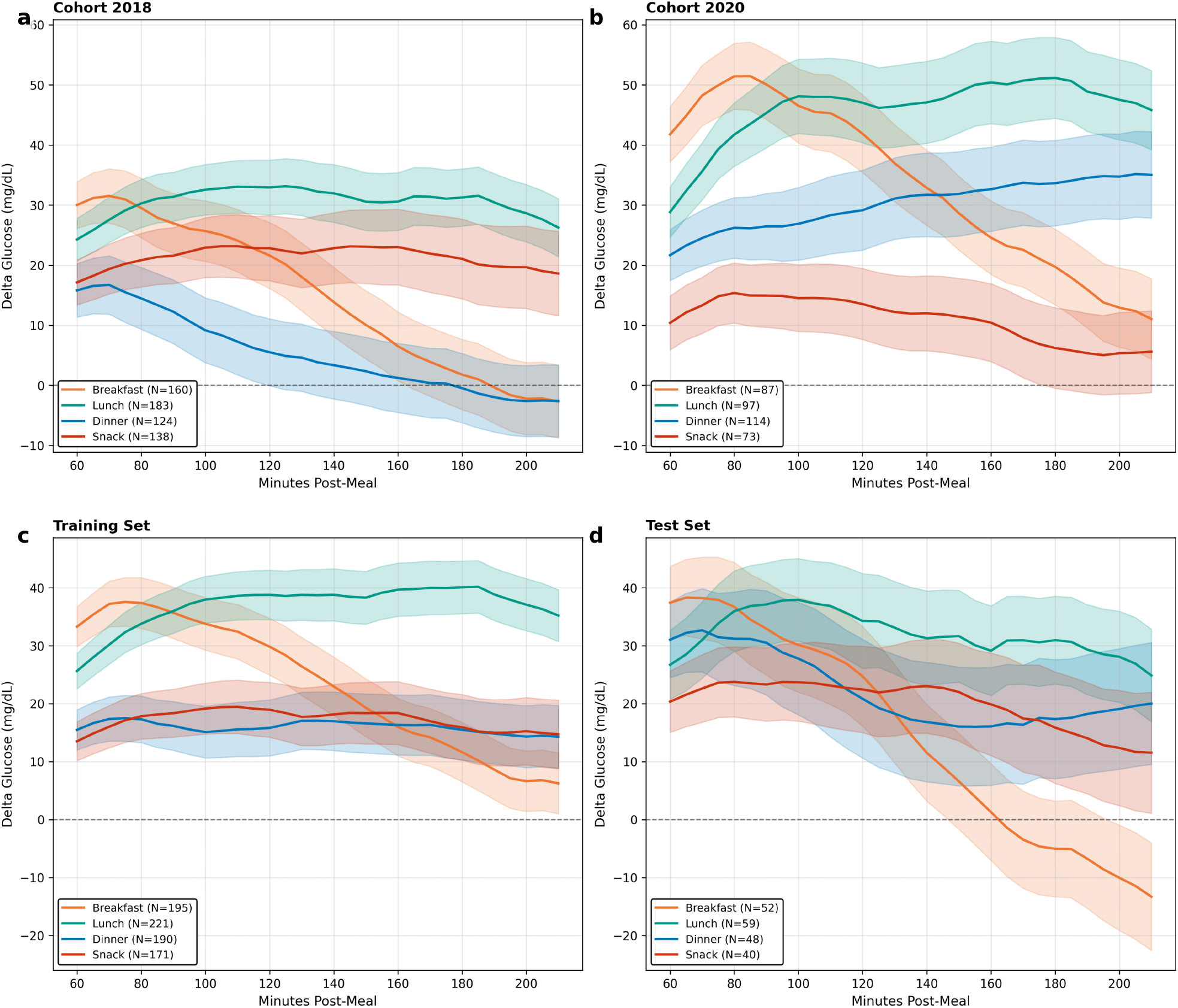
Mean postprandial delta glucose trajectories by meal type. Mean *±* 1 SE glucose excursion (Δ*G*) from 60 to 210 minutes post-meal. (**a, b**) Trajectories stratified by cohort: the 2020 cohort exhibits higher and more sustained glucose excursions than 2018, particularly at lunch and dinner, reflecting between-cohort heterogeneity in postprandial glucose control. (**c, d**) Trajectories stratified by training and test sets: qualitatively similar patterns across partitions confirm that the temporal train/test split preserves the distributional characteristics of the full dataset. Across all panels, dinner (blue) and lunch (green) show the largest and most sustained excursions, while snack (red) remains near baseline, emphasizing the meal-type heterogeneity in mediation structure reported below.

**Figure 3:**
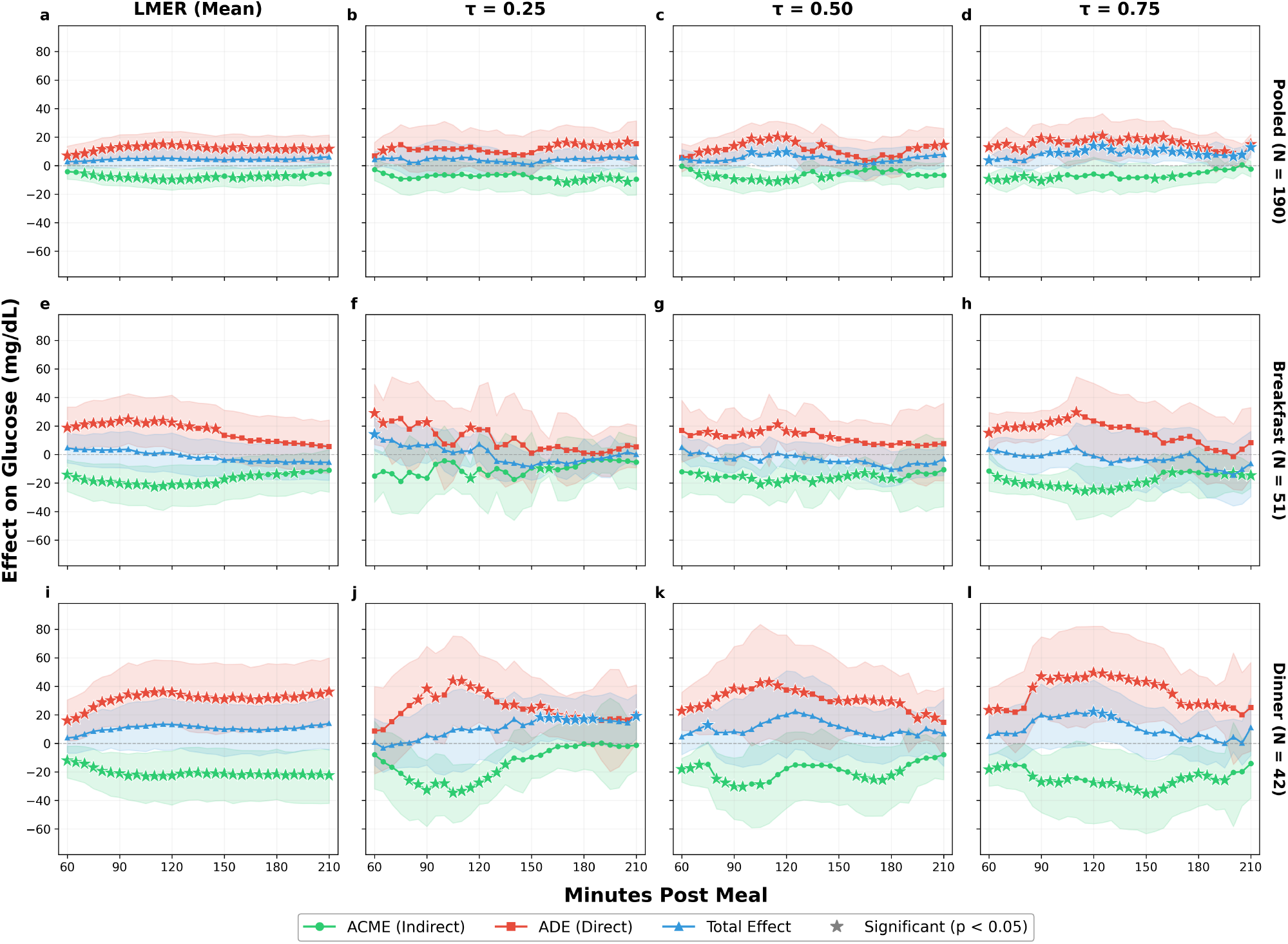
Causal mediation effects by meal type and outcome model (+30 g carbohydrate contrast). Columns show the LMER mean-level analysis and quantile regression at *τ* ∈ {0.25, 0.50, 0.75 }. Rows show pooled (all meals), breakfast, and dinner analyses. (**a–d**) At the pooled level, the non-significant LMER total effect becomes significant at *τ* = 0.50 and *τ* = 0.75, revealing distributional heterogeneity masked by mean-level analysis. (**e–h**) At breakfast, the near-cancellation of ADE and ACME persists across all quantiles, indicating well-calibrated bolusing. (**i–l**) At dinner, the ADE exceeds the ACME at every quantile, with the total effect reaching significance at *τ* = 0.75 (22.03 mg/dL, *p* = 0.04). Star markers (★) denote significance (*p <* 0.05). Green = ACME (indirect effect through insulin); Red = ADE (direct effect); Blue = Total effect.

The quantile regression analysis adds a distributional dimension to this cross-meal comparison, revealing that the mediation structure varies not only across meal types but also across quantiles of the conditional glucose response. At dinner, the incomplete insulin compensation is present across all quantiles as the ADE exceeds the ACME in magnitude at every quantile examined, and the total effect grows monotonically from 10.79 mg/dL at *τ* = 0.25 through 19.95 mg/dL at *τ* = 0.50 to 22.03 mg/dL (*p* = 0.04) at *τ* = 0.75. At breakfast, the near-cancellation of direct and mediated effects persists across all three quantiles examined, with the total effect remaining non-significant even at *τ* = 0.75 ( −1.00 mg/dL, *p* = 0.96; Table 4). This interaction implies that distributional heterogeneity in glucose control is not a uniform scaling of mean-level effects but is concentrated at specific meal types, with dinner and the upper tail of the glucose response distribution forming the locus of greatest clinical concern.

The *τ* = 0.75 analysis is suggestive of a subpopulation for whom the total carbohydrate effect is large in the upper tail despite being modest at the mean, but, as noted above, these quantile-specific total effects are exploratory and do not survive false-discovery-rate correction. The robust and FDR-corrected conclusion is that carbohydrate intake acts chiefly through a large direct effect that insulin only partially offsets; the meal- and quantile-specific patterns refine where this incomplete compensation appears largest and are offered as hypotheses for confirmatory study. If borne out, they would imply that fixed carbohydrate-to-insulin ratios implicitly targeting the average response ^28,39^ could be refined by quantile-aware and meal-type-specific adjustments for the subpopulation most at risk of postprandial hyperglycemia.

### Replication in an independent cohort

To assess generalizability and address the limited size of OhioT1DM, we replicated the full pipeline, applying identical inclusion criteria, CLAE embedding, npCBPS weighting, and mediation estimators, on the DiaTrend cohort (54 adults with type 1 diabetes; 1,890 analyzed meal events; Figure 4). The prespecified primary endpoint replicates: the pooled average direct effect of a +30 g carbohydrate contrast at 120 minutes is positive and significant (ADE = +5.3 mg/dL, 95% CI: 0.6 to 10.1, *p* = 0.02), confirming that carbohydrate intake raises postprandial glucose chiefly through a direct pathway in an independent population. Table 5 summarizes the FDR-adjusted mediation effects at the primary two-hour horizon for both cohorts: the pooled direct effect is the only effect significant in both datasets, while the OhioT1DM meal-specific effects (notably the large dinner ADE) and the DiaTrend breakfast total effect appear in single cohorts and are not jointly replicated, consistent with their exploratory status. The direct effect strengthens across the later postprandial window (ADE = +7.7 mg/dL at 180 minutes and +8.5 mg/dL at 210 minutes, both *p <* 0.001), whereas the insulin-mediated (indirect) effect is small and non-significant at every horizon (ACME between −0.2 and −1.8 mg/dL, all *p >* 0.10), exactly the direct-effect-dominant pattern observed in OhioT1DM.

**Figure 4:**
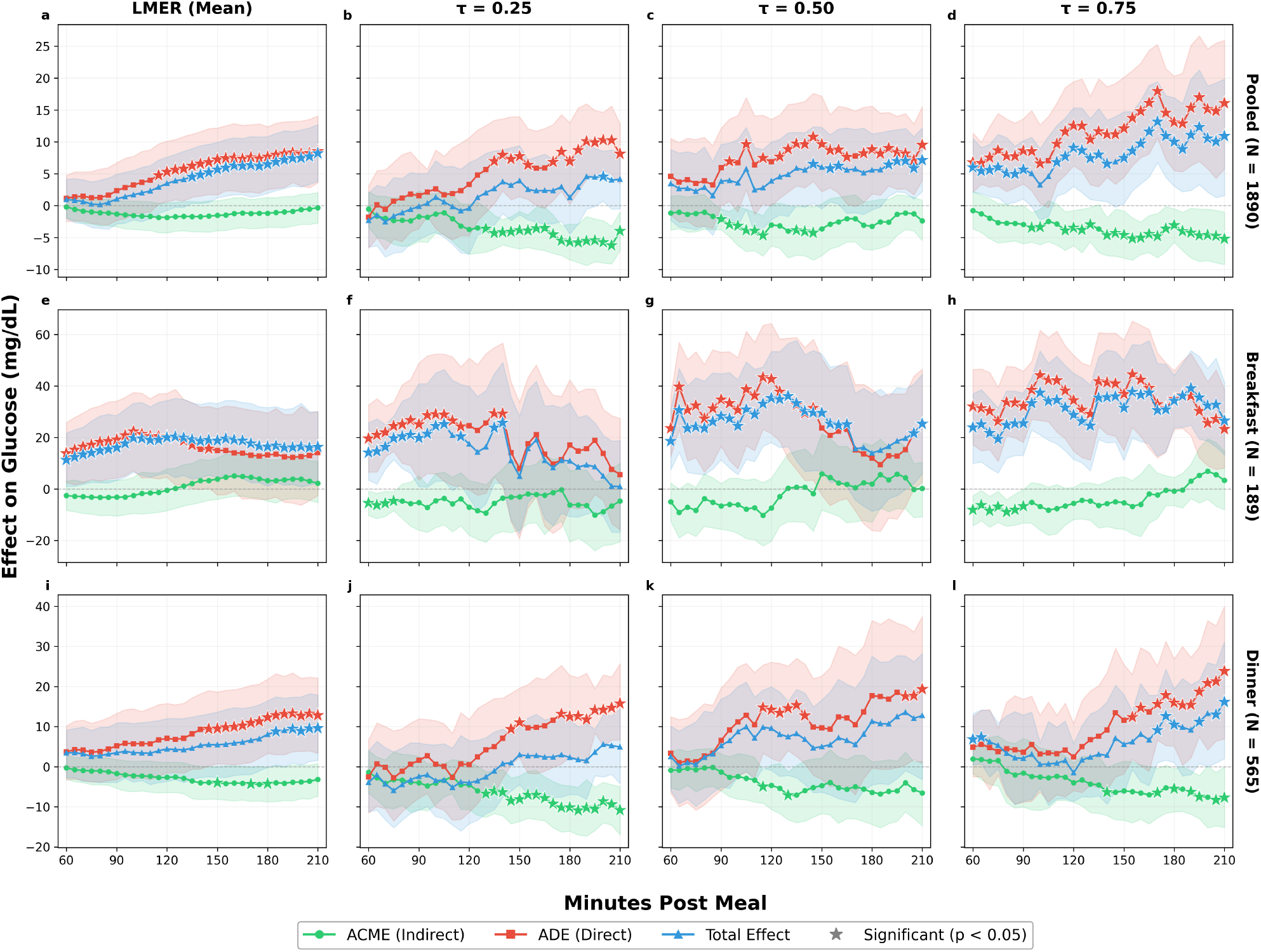
Replication of causal mediation effects in the independent DiaTrend cohort (+30 g carbohydrate contrast). Layout mirrors Figure 3: columns show the LMER mean-level analysis and quantile regression at *τ* ∈ { 0.25, 0.50, 0.75 }, and rows show pooled (all meals), breakfast, and dinner analyses (*N* = 1,890 meal events from 54 adults). The prespecified primary direct effect (pooled ADE at 120 minutes) replicates and strengthens across the postprandial window, while the insulin-mediated effect remains small and non-significant throughout. Meal- and quantile-specific effects are exploratory and are not corrected for multiplicity in this figure. Green = ACME (indirect effect through insulin); Red = ADE (direct effect); Blue = Total effect.

Consistent with the exploratory status of the OhioT1DM meal- and quantile-specific signals, the corresponding DiaTrend estimates did not reproduce as discrete, FDR-surviving effects, and the dinner-specific upper-quantile pattern was not recovered in this cohort; the direct-effect dominance, by contrast, was stable across cohorts and across a subject-cluster bootstrap (Supplementary Material). The replication therefore supports the central, multiplicity-robust conclusion of the OhioT1DM analysis while underscoring that the meal-specific timing of incomplete insulin compensation is cohort-dependent and should be treated as hypothesis-generating.

## DISCUSSION

The central contribution of this study is the demonstration that the causal pathway from carbohydrate intake to postprandial glucose is not homogeneous. It varies qualitatively across meal types and across the conditional glucose response distribution. Population-average analyses that treat insulin as a confounder or that pool across meal types obscure this heterogeneity, yielding modest and often non-significant total effects that mask clinically important patterns operating at the meal-type and distributional levels. By decomposing the total effect into direct and insulin-mediated components and estimating these components separately by meal type and outcome quantile, the present analysis reveals structure that standard approaches fail to detect.

The central, multiplicity-robust finding is that carbohydrate intake raises postprandial glucose chiefly through a large direct effect that insulin only partially offsets. The prespecified primary endpoint, the pooled average direct effect at 120 minutes, is large, survives Benjamini-Hochberg correction under every multiplicity family considered, and replicates in the independent DiaTrend cohort. Exploratory quantile analyses suggest that the net (total) effect is concentrated in the upper tail of the conditional response distribution: at the pooled level the *τ* = 0.75 total effect at 120 minutes is 13.95 mg/dL, nearly three times the mean-level estimate of 5.17 mg/dL (*p* = 0.13), because the mean averages over a distribution in which near-zero effects at lower quantiles dilute larger net increases at the upper quantiles (Figure 3). This quantile-specific total effect does not survive multiplicity correction and is reported as hypothesis-generating; if confirmed, the upper-quartile individuals it identifies would be those for whom dosing recommendations based on average effects are least adequate.

The most actionable exploratory finding concerns dinner, where both the direct and mediated effects are the largest of any meal type in absolute magnitude, but, unlike breakfast, the direct effect exceeds the mediated effect throughout the postprandial window. This imbalance is consistent with dinner bolusing being less well-calibrated to carbohydrate load than at other meals, producing a persistent net glycemic impact. The pattern appears most pronounced at *τ* = 0.75, where the dinner total effect reaches 22.03 mg/dL at 120 minutes (*p* = 0.04); however, this dinner quantile-specific total effect is exploratory and does not survive multiplicity correction, so we present incomplete insulin compensation at dinner as a hypothesis-generating signal rather than a confirmed effect. Whether it reflects suboptimal insulin-to-carbohydrate ratios at dinner, delayed carbohydrate absorption from evening meals, or reduced patient attention to bolus timing is not identifiable from these data, but the finding nominates dinner as a priority target for confirmatory study and dosing optimization. ^14,24^ These findings bear most directly on mealtime bolus decisions, which remain user-initiated in current manual and hybrid closed-loop therapy, ^28^ and on the design of future meal-aware and distribution-aware dosing algorithms. We do not claim that the dinner-specific signal is necessarily a major failure mode of fully automated post-meal correction in present-day AID; establishing this would require evaluation in contemporary AID users.

A tentative clinical implication, contingent on confirmation, is that patients whose postprandial glucose responses consistently fall in the upper quartile of the conditional distribution may benefit from more aggressive dinner bolusing than mean-level insulin-to-carbohydrate ratios prescribe. Current automated insulin delivery (AID) algorithms rely on fixed carbohydrate-to-insulin ratios that implicitly target the average response; ^28,39^ integrating quantile-aware and meal-type-specific adjustments could, if these signals are confirmed, improve glycemic outcomes for the subpopulation most at risk of postprandial hyperglycemia. ^36,40–42^

Breakfast provides a revealing contrast. The direct and insulin-mediated effects are large, individually significant, and nearly equal in magnitude, producing total effects indistinguishable from zero. This near-complete cancellation indicates that, on average, individuals in this cohort administer bolus doses that are well-calibrated to their breakfast carbohydrate intake. The magnitude of both effects, roughly double the pooled estimates, is consistent with elevated morning insulin resistance: ^38^ the same carbohydrate load produces a larger direct glycemic impact in the morning hours, but individuals compensate with correspondingly larger boluses. At *τ* = 0.75, the near-cancellation persists, reinforcing the conclusion that breakfast bolusing is well-calibrated across the glucose response distribution. From a clinical standpoint, this suggests that current breakfast bolusing practices may already be reasonably well-tuned, at least in the mean.

We emphasize that the breakfast-versus-dinner contrast is hypothesis-generating. Beyond diurnal variation in insulin sensitivity, at least four alternatives could contribute: differential carbohydrate-counting error by meal type; differences in fat, protein, and fibre that alter the timing of glucose appearance; ^16^ variation in pre-bolus timing across meals; and between-cohort heterogeneity (Figure 2). These are not separately identifiable in the present data, and we do not claim a single mechanism.

From a methodological standpoint, the CLAE approach addresses a practical challenge in observational causal inference: how to adjust for high-dimensional pre-treatment confounders when hand-crafted feature engineering is infeasible and sample sizes are modest. Unlike standard autoencoders that optimize solely for reconstruction, ^33,43,44^ the CLAE adds balance- and linearity-oriented penalties to the training objective to improve adjustment for measured pre-meal confounders. ^34,45^ We frame CLAE as a compact, balance-oriented confounder-adjustment representation for the high-dimensional pre-meal trajectory in a small-sample setting, rather than as a method that outperforms simpler adjustments on prediction or that establishes the identification assumptions; the reported analyses use principal components of this representation as covariates.

A natural concern with using learned embeddings in causal inference is interpretability. We emphasize that φ serves exclusively as a confounder adjustment variable, not as a quantity of direct scientific interest. The causal estimands (ACME, ADE) are defined in terms of carbohydrate intake and insulin dose, both of which remain fully interpretable on their original clinical scales. The role of φ is analogous to including a flexible function of baseline covariates in a propensity score model. ^46^ Furthermore, because φ is derived exclusively from pre-treatment data (the [ −120, 0) minute window), it cannot introduce post-treatment bias or encode information about the treatment, mediator, or outcome.

The ablation study provides empirical guidance on which constraints matter, and we correct a reporting error in an earlier version of this manuscript. The embedding’s intrinsic covariate balance is achieved primarily through the encoder architecture and the linearization penalty; the marginal contribution of the balancing penalty to the intrinsic balance score is negligible to negative (Supplementary Tables 13–16). The selected configuration was therefore chosen not for its intrinsic balance score but for the npCBPS-weighted covariate balance it achieves in the mediation sample. In practice, retaining three principal components from the CLAE embeddings eliminated multicollinearity, and npCBPS weighting on these components reduced the mean absolute correlation with treatment by 97.2% (weighted balance 0.961), with an effective sample size of 95.5%, confirming that adequate balance on measured covariates was achieved without reliance on extreme weights. ^47^

Because the no-unmeasured-confounding component of sequential ignorability is untestable, we quantified the robustness of the mediation conclusions to its violation using the sensitivity parameter *ρ* of Imai et al., ^29^ the residual mediator-outcome correlation at which the estimated indirect effect would vanish. The analysis indicates that the estimated insulin-mediated effect is nullified by only a small degree of unmeasured mediator-outcome confounding (*ρ*^∗^≈ −0.05). The indirect (insulin-mediated) pathway should therefore be interpreted cautiously. The direct-effect conclusion, by contrast, is robust to multiple-testing correction and replicates in an independent cohort, and does not depend on the mediator-outcome ignorability condition to the same degree.

Several limitations qualify these findings. The combined 2018 and 2020 OhioT1DM cohorts ^48,49^ provide rich longitudinal data but comprise a modest number of subjects, limiting generalizability to broader T1DM populations. The meal-type-stratified analyses yield sample sizes of 51 (breakfast), 58 (lunch), 42 (dinner), and 39 (snack) observations in the test set.

Dinner glucose trajectories also differ between the 2018 and 2020 cohorts (Figure 2a,b). Because each OhioT1DM cohort contains only six subjects, fully within-cohort mediation estimates are imprecise; we therefore treat the meal-stratified results as exploratory and rely on the independent DiaTrend cohort for cross-population robustness, in which the direct-effect finding replicates while its meal-specific timing differs.

Carbohydrate intake is self-reported and subject to counting error that is plausibly differential across meal types: dinners are more often mixed, home-cooked meals and may be estimated less accurately than packaged or repetitive breakfasts and snacks. ^50^ Whereas non-differential error attenuates estimates toward the null, ^51^ systematic under-reporting at dinner would inflate the estimated direct effect per reported gram. A bias analysis indicates that approximately 37% systematic dinner under-reporting, relative to breakfast, would be required to account for the observed breakfast-versus-dinner direct-effect ratio by measurement error alone; carbohydrate underestimation of this kind is common (63% of meals; approximately 21% on average), ^50^ so this alternative cannot be excluded. We therefore present the breakfast-versus-dinner contrast as hypothesis-generating.

Meal logs in both cohorts record carbohydrate amount only; fat, protein, fibre, and glycemic index, which are established modifiers of postprandial glucose appearance, ^16^ were unavailable and could not be adjusted for. Their omission is a limitation and a plausible contributor to meal-type differences.

Mediation was not detected at lunch or snack; full results for these meal types appear in the Supplementary Material. These null findings likely reflect substantive differences in meal-related insulin dosing behavior rather than an absence of causal structure. ^52^ Snacking is inherently more heterogeneous than structured meals, with greater variability in carbohydrate content, timing, and whether a bolus is administered at all. Approximately 50% of snack boluses are missed in youth with T1DM, ^53^ and unannounced snack intake degrades glucose control even under automated insulin delivery. ^54^ At lunch, insulin-to-carbohydrate coupling may be more moderate and consistent than at other meals, producing neither the amplified opposing pathways seen at breakfast nor the large, incompletely offset effects seen at dinner. More broadly, breakfast, lunch, and dinner differ in their postprandial glucose profiles, ^55^ with diurnal variation in insulin sensitivity further modulating the treatment-mediator-outcome relationship across meal types. ^38^ Larger datasets with more subjects and greater meal-type coverage would help determine whether these explanations fully account for the null findings or whether moderate effects exist below the detection threshold of the present analysis.

Looking ahead, the heterogeneity documented in this study, varying across subjects, meal types, and quantiles of the conditional glucose distribution, suggests several directions for extending the causal mediation framework to personalized diabetes management.

The most immediate extension is transfer learning. ^56,57^ The per-stratum sample sizes in our meal-type analyses (42–58 test observations) limit the precision of subject-specific or meal-type-specific mediation estimates. Pretraining CLAE-type architectures on pooled or external CGM datasets and fine-tuning on individual patients or meal types would allow the estimation of personalized mediation effects that our current sample size cannot support. This strategy has proven effective in related clinical domains. Convolutional networks pre-trained on large ECG datasets achieve cardiologist-level arrhythmia detection when transferred to smaller cohorts, ^58^ and deep transfer learning combined with data augmentation has been shown to improve glucose prediction in type 2 diabetes patients. ^59^ Richer temporal architectures, such as the convolutional-recurrent models proposed for glucose forecasting, ^60,61^ could serve as stronger approximate encoders in a transfer learning pipeline, capturing nonlinear dynamics in pre-meal trajectories that the linear CLAE encoder may not fully represent.

The single most important next step, however, is external validation in larger and more diverse cohorts, and ideally in contemporary automated-insulin-delivery users. We have taken a first step here by replicating the primary direct-effect finding in the independent DiaTrend cohort, but confirmatory testing of the exploratory meal- and quantile-specific signals will require prospectively larger, demographically broader data. We explicitly do not advocate data augmentation ^62,63^ as a remedy for the present sample size: augmenting within twelve subjects cannot recover generalization error and risks entrenching artefacts of the current sample. External-cohort validation and transfer learning are the appropriate responses to this data limitation, and we prioritize them accordingly.

Finally, our quantile regression results highlight that the clinically critical effects concentrate in the upper tail of the glucose response distribution, a regime that is inherently underrepresented in mean-focused analyses. This connects to the broader literature on learning from imbalanced distributions, ^64–66^ where standard loss functions and sampling procedures systematically underweight the tails. Integrating distributional reweighting or quantile-focused loss functions into the autoencoder training objective could improve representation quality precisely where it matters most for clinical decision-making. More broadly, the growing availability of large-scale CGM datasets ^67,68^ and the maturation of machine learning methods for diabetes research create favorable conditions for scaling the causal mediation approach beyond the single-cohort analysis presented here.

## METHODS

### Datasets

We analyzed the OhioT1DM 2018 and 2020 cohorts, comprising twelve adults with type 1 diabetes monitored over eight weeks each. ^48,49,69^ Participants used Medtronic 530G or 670G insulin pumps paired with Enlite CGM sensors recording interstitial glucose at 5-minute intervals. ^35,70,71^ Physiological signals (heart rate, skin temperature, step counts) were collected via wrist-worn fitness bands. Meal events with carbohydrate estimates, bolus insulin deliveries, and basal rates were self-reported or logged by the pump. ^19^ All analyses use de-identified data in accordance with the dataset’s license agreement. For each subject, the first 65% of days (chronologically) were assigned to training and the final 35% to testing, ensuring strict temporal separation. Standardization parameters were computed on training data only.

To assess generalizability and address the limited size of OhioT1DM, we additionally analyze DiaTrend ^72^, an independent type 1 diabetes cohort, applying identical inclusion criteria and analysis. DiaTrend was selected because it provides the synchronized meal-level carbohydrate, bolus-insulin, and high-frequency CGM records required for the present estimands, which are not available at this granularity in registry datasets such as T1D Exchange or in predominantly non-type-1 CGM foundation cohorts such as GluFormer ^73^. We analyze 54 adults with type 1 diabetes spanning the dataset’s two device cohorts; meal windows, treatment, mediator, and outcome are defined exactly as for OhioT1DM, with an analogous within-subject temporal split in which the final portion of each subject’s record is held out for testing. After identical filtering, the heldout test set comprised 1,890 demographics-complete meal events, on which the replication analysis was performed (Supplementary Material). DiaTrend is available through the Sage Bionetworks Synapse platform to certified users under its Terms of Use; its access terms are less restrictive than those governing the OhioT1DM data.

### Data preprocessing and defining meal-centered windows

Missing CGM values were imputed via linear interpolation when fewer than six consecutive readings were absent. ^2,68,74^ For each logged meal, we constructed a window spanning [ −120, 0) minutes (pre-meal) through +210 minutes (post-meal) on the 5-minute CGM grid. Windows with overlapping meals (*<*90 minutes apart), excessive missing data, or unresolved CGM artifacts were excluded; approximately 58 observations with post-mediator-window boluses were also excluded to preserve temporal ordering. After filtering, the analytic sample comprised 976 meal windows (777 training, 199 test). Table 1 summarizes treatment, mediator, and pre-meal glucose distributions by cohort and meal type.

### Treatment, mediator, and outcome

The treatment *Z*_*i*_ is the self-reported carbohydrate content (grams) of meal *i*.^16,20^ Treatment contrasts are defined relative to meal-type-specific medians, with the primary contrast set at +30 g. The mediator *M*_*i*_ is the total bolus insulin (units) aggregated over [ −120, +60] minutes relative to the meal, capturing pre-meal, meal-time, and early correction boluses. ^24,25^ This window terminates at +60 minutes to ensure the mediator is fully determined before the first outcome horizon. Approximately 12% of observations have zero bolus insulin. The outcome Δ*G*_*i*_(*t*) = *G*_*i*_(*t*) −*G*_*i*_(0) is the glucose excursion from the meal-time baseline, evaluated at *t* ∈ {60, 90, 120, 150, 180, 210 } minutes postmeal. ^15,42^

### Causally-constrained linear autoencoder

Causal mediation analysis requires adjustment for confounders of the treatment-mediator and mediator-outcome relationships. ^29^ In our setting, these confounders are contained in longitudinal pre-meal CGM trajectories (24 *×* 5 = 120 features per observation), which cannot serve as covariates directly given the sample size. We developed a Causally-constrained Linear Autoencoder (CLAE) that compresses pre-meal time series into low-dimensional embeddings φ ∈ ℝ ^8^ while incorporating constraints designed to improve covariate balance and adjustment for measured pre-meal confounders. ^45,75^

The encoder receives five input channels (glucose, step counts, basal insulin, meal carbohydrates, heart rate) from the [ −120, 0) pre-meal window; bolus insulin is excluded to prevent mediator leakage into the learned confounders. We evaluated both convolutional (CNN) and recurrent (LSTM) encoder architectures. The CNN encoder applies successive convolutional blocks with increasing filter depth (32, 64, 128 filters) followed by global average pooling. The LSTM encoder processes the temporal sequence through a 64-unit recurrent layer. Both architectures concatenate the encoded representation with a summary of temporal feature means before a final linear projection to 8-dimensional embeddings. Categorical features (meal type, subject ID) were excluded from the encoder because they strongly correlate with treatment and caused poor balance in learned representations. Architecture details, including layer specifications and regularization parameters, are provided in Table 2.

**Table 2:**
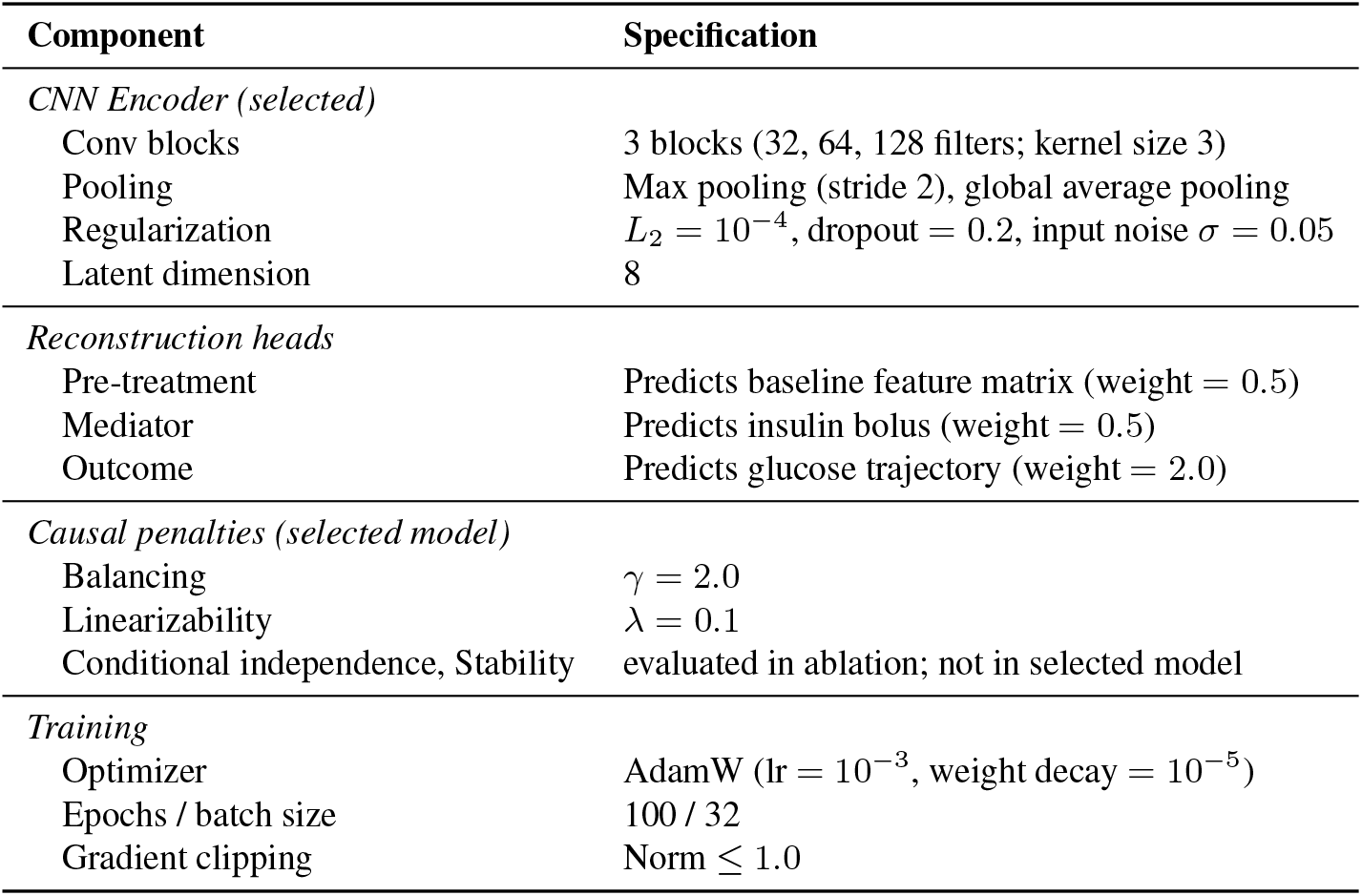
CLAE architecture and training configuration.

| Component | Specification |
| --- | --- |
| *CNN Encoder (selected)* |  |
| Conv blocks | 3 blocks (32, 64, 128 filters; kernel size 3) |
| Pooling | Max pooling (stride 2), global average pooling |
| Regularization | $L_2 = 10^{-4}$, dropout = 0.2, input noise $\sigma = 0.05$ |
| Latent dimension | 8 |
| *Reconstruction heads* |  |
| Pre-treatment | Predicts baseline feature matrix (weight = 0.5) |
| Mediator | Predicts insulin bolus (weight = 0.5) |
| Outcome | Predicts glucose trajectory (weight = 2.0) |
| *Causal penalties (selected model)* |  |
| Balancing | $\gamma = 2.0$ |
| Linearizability | $\lambda = 0.1$ |
| Conditional independence, Stability | evaluated in ablation; not in selected model |
| *Training* |  |
| Optimizer | AdamW (lr = $10^{-3}$, weight decay = $10^{-5}$) |
| Epochs / batch size | 100 / 32 |
| Gradient clipping | Norm $\leq 1.0$ |

**Table 3:**
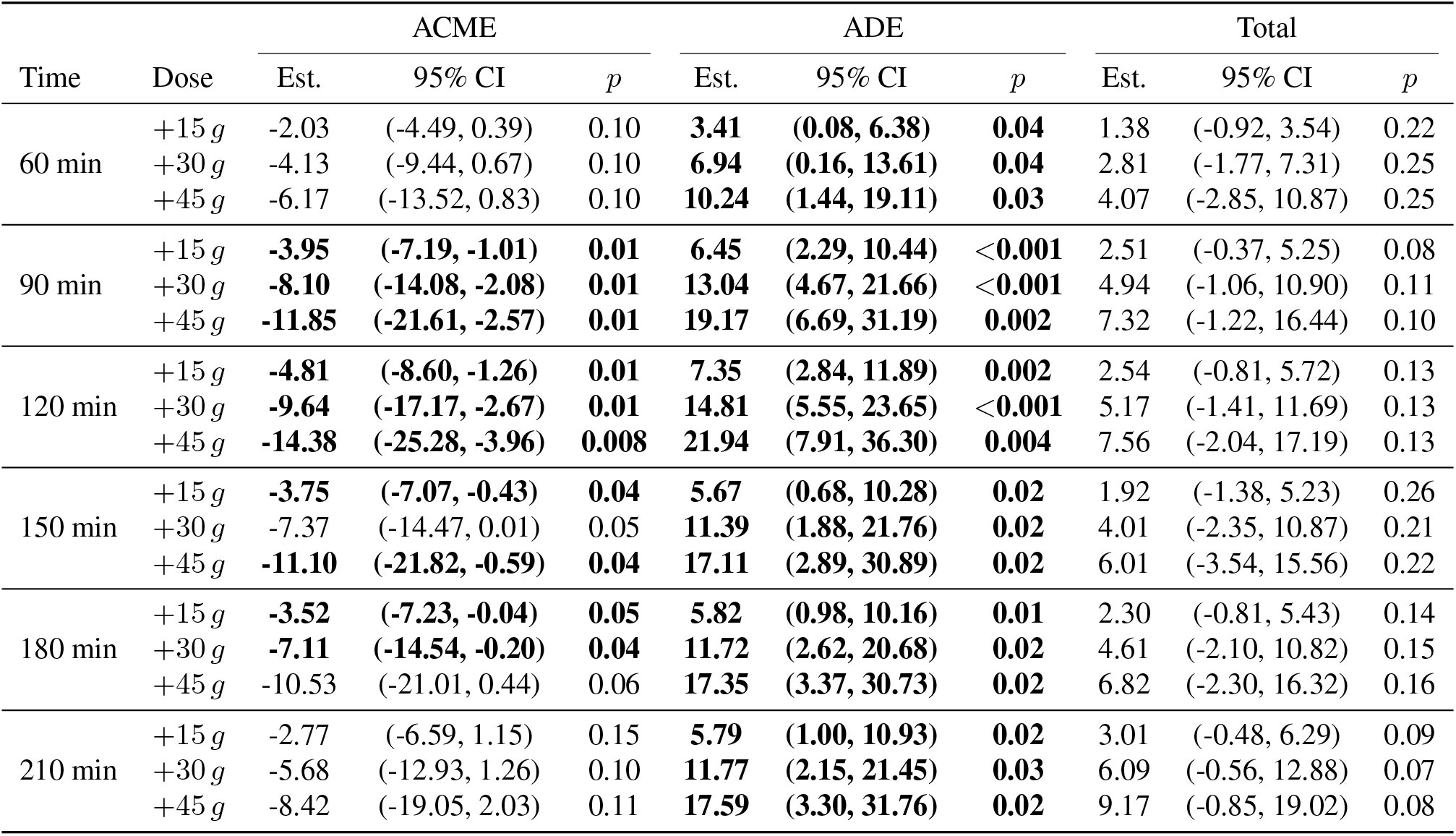
PCA covariate — Pooled (all meals): LMER causal mediation effects across treatment doses (*N* = 190 meal observations, all contrast sizes). Time = postprandial measurement time point in minutes after meal start. Dose = hypothetical increase in carbohydrate intake (grams) above the meal-type-specific median. ACME = Average Causal Mediation Effect (indirect effect mediated through insulin); ADE = Average Direct Effect (effect not mediated through insulin); Total = ACME + ADE. Est. = point estimate (mg/dL); 95% CI from quasi-Bayesian approximation with 1000 Monte Carlo simulations; *p* = two-sided p-value testing the null hypothesis that the effect equals zero. Significant results (*p <* 0.05) are shown in bold.

**Table 4:**
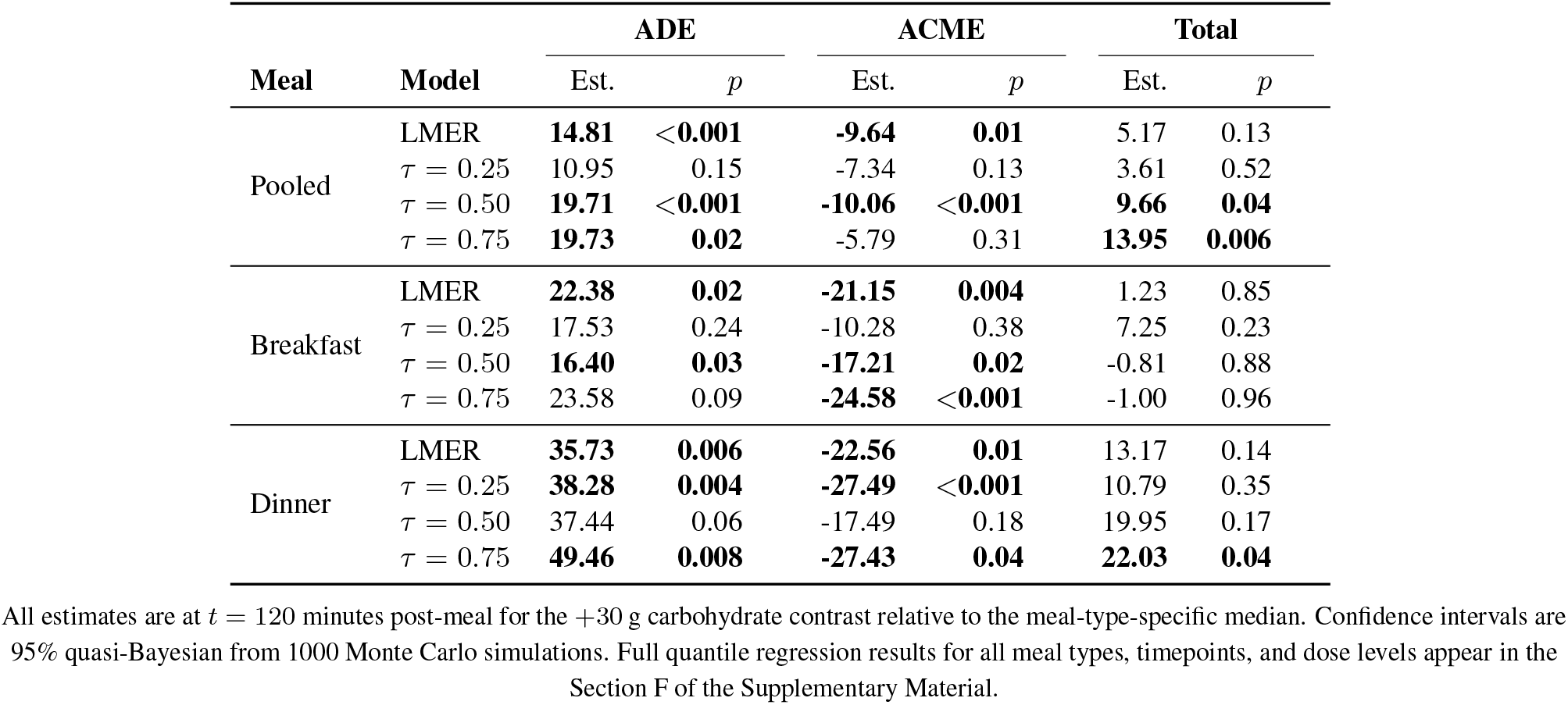
Summary of mediation effects at 120 minutes post-meal, pooled across meal types (+30 g carbohydrate contrast, *N* = 190 meal observations). Entries report point estimates (mg/dL) and *p*-values for the average direct effect (ADE), average causal mediation effect (ACME), and total effect at three quantiles of the conditional glucose response distribution. The LMER meanlevel estimate is included for comparison. Boldface indicates statistical significance (*p <* 0.05).

| Meal | Model | ADE |  | ACME |  | Total |  |
| --- | --- | --- | --- | --- | --- | --- | --- |
| | | Est. | $p$ | Est. | $p$ | Est. | $p$ |
| Pooled | LMER | **14.81** | **<0.001** | **-9.64** | **0.01** | 5.17 | 0.13 |
| | $\tau = 0.25$ | 10.95 | 0.15 | -7.34 | 0.13 | 3.61 | 0.52 |
| | $\tau = 0.50$ | **19.71** | **<0.001** | **-10.06** | **<0.001** | **9.66** | **0.04** |
| | $\tau = 0.75$ | **19.73** | **0.02** | -5.79 | 0.31 | **13.95** | **0.006** |
| Breakfast | LMER | **22.38** | **0.02** | **-21.15** | **0.004** | 1.23 | 0.85 |
| | $\tau = 0.25$ | 17.53 | 0.24 | -10.28 | 0.38 | 7.25 | 0.23 |
| | $\tau = 0.50$ | **16.40** | **0.03** | **-17.21** | **0.02** | -0.81 | 0.88 |
| | $\tau = 0.75$ | 23.58 | 0.09 | **-24.58** | **<0.001** | -1.00 | 0.96 |
| Dinner | LMER | **35.73** | **0.006** | **-22.56** | **0.01** | 13.17 | 0.14 |
| | $\tau = 0.25$ | **38.28** | **0.004** | **-27.49** | **<0.001** | 10.79 | 0.35 |
| | $\tau = 0.50$ | 37.44 | 0.06 | -17.49 | 0.18 | 19.95 | 0.17 |
| | $\tau = 0.75$ | **49.46** | **0.008** | **-27.43** | **0.04** | **22.03** | **0.04** |

All estimates are at $t = 120$ minutes post-meal for the +30 g carbohydrate contrast relative to the meal-type-specific median. Confidence intervals are 95% quasi-Bayesian from 1000 Monte Carlo simulations. Full quantile regression results for all meal types, timepoints, and dose levels appear in the Section F of the Supplementary Material.

**Table 5:**
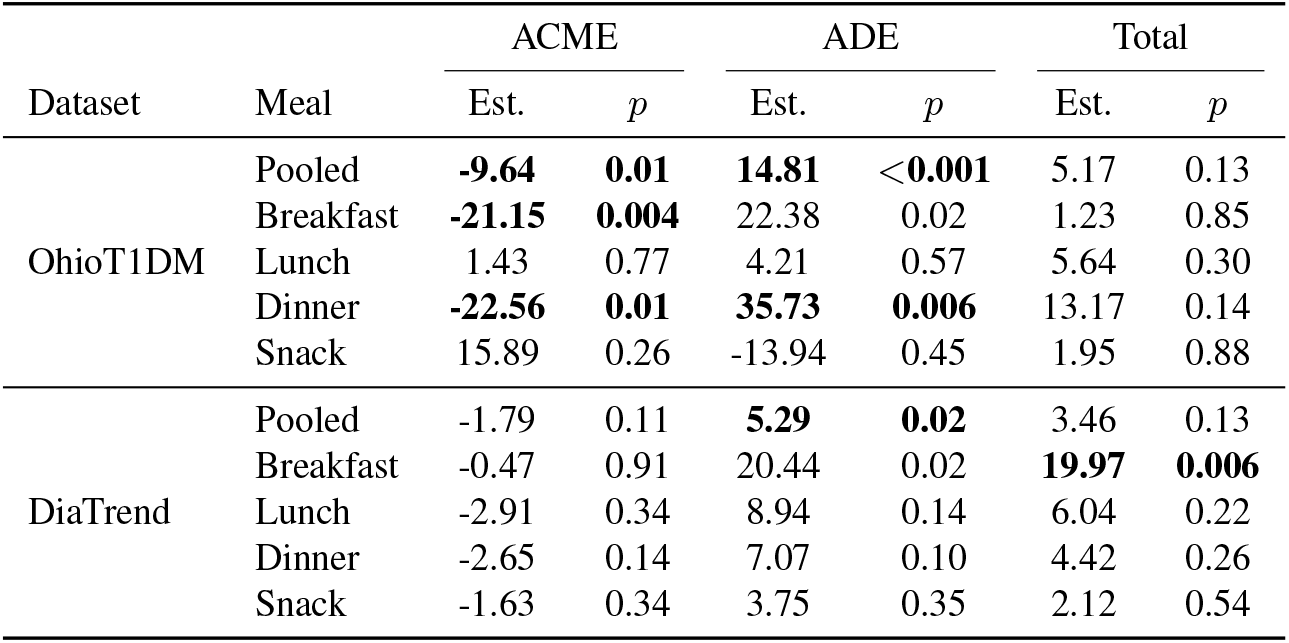
FDR-adjusted causal mediation effects at the two-hour (120 min) postprandial horizon for a +30 g carbohydrate contrast (OhioT1DM, *N* = 190; DiaTrend, *N* = 1,890 test meal observations). Mean-level (LMER) average causal mediation effect (ACME, through insulin), average direct effect (ADE), and total effect by meal stratum. Est. = point estimate (mg/dL); *p* from quasi-Bayesian approximation with 1000 Monte Carlo simulations. The pooled ADE is the prespecified primary endpoint (tested at *α* = 0.05); all other effects are secondary, and values in boldface are significant after Benjamini–Hochberg false-discovery-rate control within the meal stratum.

The training objective combines two components: reconstruction losses that ensure the embeddings φ retain clinically relevant information, and causal regularization penalties that encourage the embeddings to satisfy the conditions required for valid mediation. ^76^

The reconstruction component includes four prediction heads, each of which receives φ as input and predicts a different target: the pre-meal feature matrix (ensuring φ captures baseline physiological state), the insulin bolus (ensuring φ contains mediator-relevant information), and the postprandial glucose trajectory (ensuring φ is informative for the primary outcome). ^43^ A fourth head predicting treatment assignment is included in the architecture for ablation purposes but is assigned zero weight in the final configuration. Assigning it positive weight would conflict with the balancing penalty described below, because an encoder that accurately predicts treatment necessarily learns representations that distinguish higher-from lower-carbohydrate meals, undermining covariate balance. ^46^ Loss weights prioritize outcome prediction (weight = 2.0) over pretreatment reconstruction and mediator prediction (each = 0.5), reflecting the relative importance of these targets for the downstream causal estimands.

The causal regularization component comprises up to four penalties (Table 2); the configuration used for all reported analyses retains the balancing and linearizability penalties, while the conditional independence and stability penalties were evaluated in the ablation but not retained (see Results). The balancing penalty penalizes distributional differences in the embeddings between higher- and lower-carbohydrate meals, encouraging the learned representation to be less predictive of treatment and thereby improving covariate balance. ^77,78^ The linearizability penalty encourages approximate linearity between φ and the outcome, ensuring compatibility with the linear models used in the mediation analysis. ^32^ The conditional independence penalty targets the core identification assumption of mediation analysis (sequential ignorability; see below) by penalizing residual association between treatment and mediator after adjusting for φ. ^29,79^ The stability penalty discourages sensitivity of the embedding covariance structure to resampling. Before applying these penalties, the embeddings pass through a learned nonlinear basis expansion with a residual connection, allowing the model to accommodate nonlinear relationships without requiring the user to specify a fixed functional form. ^80,81^

An ablation study over five penalty configurations and both architectures, each trained with three random seeds, was used to select the final model. We prioritized covariate balance subject to adequate outcome prediction on the heldout test set (Supplementary Tables 13–16). After encoding, PCA (fit on training data only) reduces the embeddings to orthogonal components; the first 3 PCs (capturing ∼ 90% of latent variance) serve as covariates in the mediation mod-els ^34,82^

### Causal identification

Causal identification of mediation effects requires the sequential ignorability assumption. ^29,30^ This assumption comprises two conditions. The first requires that, conditional on pre-meal state (represented by φ), the carbohydrate content of a meal is effectively randomly assigned with respect to subsequent glucose and insulin responses. The second requires that, conditional on both carbohydrate intake and pre-meal state, the insulin bolus dose is effectively randomly assigned with respect to the glucose outcome. ^79^

Formally, let *Y* (*z, m*) denote the potential outcome under treatment level *z* and mediator level *m*, and let *M* (*z*) denote the potential mediator under treatment *z*. Sequential ignorability requires:

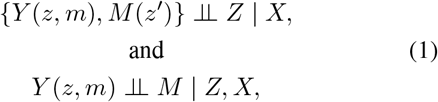

where *X* denotes the principal components of φ. The CLAE penalties are designed to reduce residual association between treatment and mediator that is attributable to measured pre-meal trajectories. They do not, and cannot, guarantee the sequential-ignorability assumptions, which require no unmeasured confounding of the treatment, mediator, and outcome, an assumption that is untestable in observational data. We therefore interpret the estimated effects as covariate-adjusted associations under an explicitly stated identification assumption, and we assess their robustness to violation of that assumption below.

### Confounder balancing and causal mediation analysis

We estimate balancing weights using non-parametric covariate balancing propensity scores (npCBPS) for continuous treatments, ^83^ regressing carbohydrate intake on the first 6 PCs of φ, glucose at meal time, and a cohort indicator. Glucose at meal time and cohort are included in the balancing model but excluded from the mediation model formulas to avoid collider bias.

For each outcome horizon, we decompose the total effect of carbohydrate intake on glucose response into the average causal mediation effect (ACME, operating through insulin), the average direct effect (ADE, all other pathways), and their sum (total effect) using the framework of Imai et al. ^29,30^ Effects are estimated at three treatment contrasts: +15, +30, and +45 g of carbohydrates above the meal-type-specific median. These contrasts represent clinically interpretable increments, roughly equivalent to adding a small snack (+15 g), a slice of bread or medium fruit (+30 g), or a substantial side dish (+45 g) to a typical meal. ^84^ By anchoring contrasts at the meal-type-specific median rather than a global threshold, we account for the substantial differences in typical carbohydrate intake across breakfast, lunch, dinner, and snack. The mediator model is a left-censored Tobit regression of insulin bolus on treatment and PCs to account for zero-inflation. ^85^ For the outcome model, we employ two complementary specifications. The primary mean-level specification is a linear mixed-effects regression (LMER) of Δ*G*_*i*_(*t*) on treatment, mediator, and PCs, with a random intercept for subject to account for within-subject correlation across repeated meal observations. To characterize distributional heterogeneity in treatment effects, we additionally estimate the outcome model via quantile regression (QR) at *τ* ∈ { 0.25, 0.50, 0.75}, which models the conditional quantiles of the glucose response distribution rather than the conditional mean. ^86,87^ Both outcome specifications are weighted by npCBPS weights and paired with the same Tobit mediator model. ^77^ Mediation effects are estimated via quasi-Bayesian approximation with 1000 Monte Carlo simulations using the `mediate` package in R. ^32,88^

### Statistical inference and multiplicity

We prespecified a single primary endpoint, the pooled mean-level average direct effect of a +30 g carbohydrate contrast at 120 minutes post-meal, tested at *α* = 0.05; 120 minutes corresponds to the guideline-standard two-hour postprandial endpoint. ^89,90^ All other reported effects are secondary and exploratory; we control the false discovery rate using the Benjamini-Hochberg procedure ^91^ within each meal stratum, and additionally report pooled and full-grid corrections as sensitivity analyses. Postprandial time-courses are interpreted descriptively. Nominal, uncorrected *p*-values for exploratory analyses are reported as hypothesis-generating. To probe robustness of the mediation conclusions to unmeasured mediator-outcome confounding, we report the sensitivity parameter *ρ* of Imai et al. ^29^, the residual mediator-outcome correlation at which the estimated indirect effect would be nullified.

### Model validation

Architecture and penalty configuration were selected on held-out test set performance, averaged across three random seeds. Embedding quality was assessed via outcome *R*^2^, mediator *R*^2^, covariate balance score, and inrange AUC for the clinically relevant 70–180 mg/dL glucose range. ^2^

### Ethics

This study is a secondary analysis of two previously collected, fully de-identified type 1 diabetes datasets; the authors collected no new data and had no contact with human participants. The OhioT1DM data were collected from participants who provided written informed consent for their de-identified data to be shared for research use ^48,49^; these data were obtained under a Data Use Agreement with Ohio University (OU Ref. D201804), and the University of California, Irvine determined that their secondary analysis did not require Institutional Review Board review. The DiaTrend data were collected under two studies approved by the Dartmouth College Committee for the Protection of Human Subjects (protocols STUDY00031632 and STUDY00023559); all participants provided verbal and written informed consent, including consent to share their data openly with the research community, and these data were accessed through the Sage Bionetworks Synapse platform under its Terms of Use. ^72^ Both source studies were conducted in accordance with the principles of the Declaration of Helsinki.

## DATA AVAILABILITY

The data that support the findings of this study are from the 2018 and 2020 Ohio T1DM cohort ^48,49^. These data are held by the dataset custodians (Ohio University) and were used under a data use agreement (DUA); therefore, restrictions apply, and the raw records are not publicly available. The data are, however, available upon request and the completion of a DUA with Ohio University. External validation used the DiaTrend dataset ^72^, which is available through the Sage Bionetworks Synapse platform (https://www.synapse.org) to certified users following acceptance of its Terms of Use and approval of a Data Access Request; its access terms are less restrictive than those governing the OhioT1DM data.

## CODE AVAILABILITY

The code is available in a GitHub repository: https://github.com/spencha/causal-mediation-cgm.

## ACKNOWLEDGEMENTS

The authors thank Dr. Jiuchen Zhang for early discussions that suggested causal mediation analysis as a methodological direction. We acknowledge the research participants who contributed data and the DiaTrend study investigators ^72^ for making the validation dataset available. This research is supported by NIH 1T32GM152338-01A1 and NCI 1R01CA297869.

## AUTHOR CONTRIBUTIONS

SH formulated the problem, developed the methodology, implemented the computer code, performed all computations and analyses, and wrote the manuscript. AQ conceptualized the core idea of the topic, supervised the work and provided critical feedback on methodology and interpretation.

