## Supplementary Materials for "Causal Mediation Pathways in Continuous Postprandial Glucose Monitoring for Type 1 Diabetes Patients"

Spencer Hilligoss and Annie Qu

July 10, 2026

#### A Data Distribution

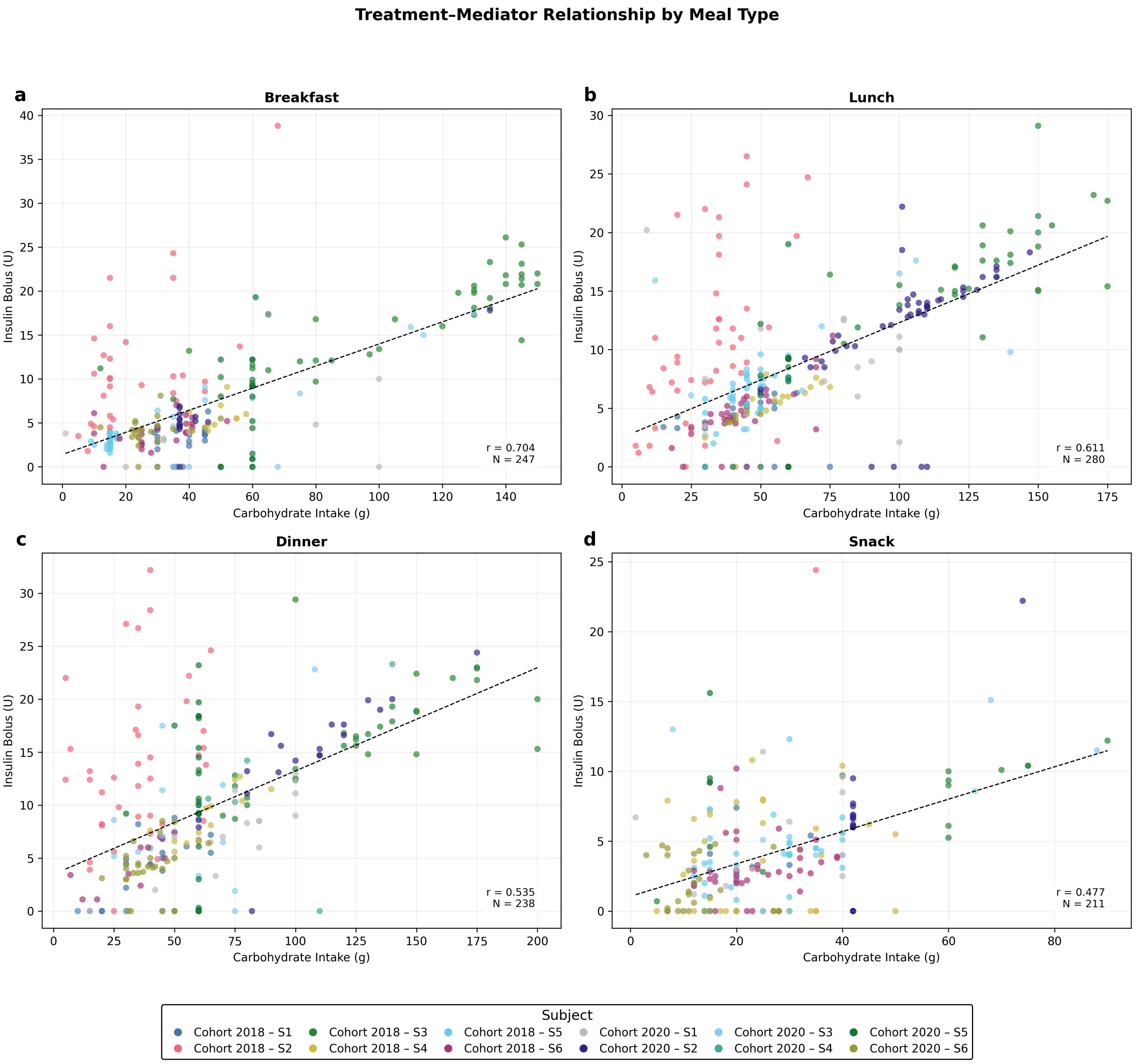


**Supplementary Figure 1: Treatment-mediator relationship by meal type.** Scatter plots of carbohydrate intake (treatment) versus insulin bolus (mediator) for each meal type.

**Supplementary Table 1: Data characteristics by data split and meal type.** Summary statistics for carbohydrate intake (treatment), insulin bolus (mediator), and pre-meal glucose stratified by training/test split and meal category. N = number of meal observations used in the analysis; SD = standard deviation; values are mean ± SD; % Zero Bolus = percentage of meals with no insulin bolus.

| | | | Carbs (g) | | Bolus (U) | | % Zero | Glucose (mg/dL) |
|---|---|---|---|---|---|---|---|---|
| Split | Meal Type | N | Mean | SD | Mean | SD | Bolus | Mean ± SD |
| Test | Breakfast | 52 | 49.5 | 36.0 | 7.77 | 6.23 | 3.8 | 175.1 ± 66.8 |
| | Lunch | 59 | 65.0 | 41.7 | 9.70 | 6.43 | 6.8 | 147.8 ± 58.8 |
| | Dinner | 48 | 67.3 | 40.9 | 9.62 | 7.07 | 12.5 | 149.6 ± 54.3 |
| | Snack | 40 | 28.1 | 16.9 | 4.12 | 4.22 | 20.0 | 142.3 ± 46.5 |
| Train | Breakfast | 195 | 46.1 | 35.5 | 7.16 | 6.38 | 8.7 | 158.1 ± 54.5 |
| | Lunch | 221 | 64.2 | 36.0 | 8.58 | 5.83 | 9.0 | 141.2 ± 47.9 |
| | Dinner | 190 | 62.2 | 37.1 | 9.66 | 6.88 | 11.6 | 155.8 ± 53.6 |
| | Snack | 171 | 26.6 | 15.7 | 4.19 | 3.77 | 20.5 | 143.8 ± 59.7 |

### B npCBPS Balancing

After applying non-parametric covariate-balancing propensity score (npCBPS) weights, all four covariates ($PC_1$, $PC_2$, $PC_3$, and standardized glucose at meal) achieved balance, with correlations with treatment reduced to $|r| < 0.01$. The mean absolute correlation with treatment dropped from 0.082 to 0.002, a 97.2% reduction. The most imbalanced covariate prior to weighting ($PC_3$, $r = -0.121$) was reduced to $r < 0.001$ after weighting. The effective sample size after weighting was 190 of 199 observations (95.5%), indicating that balance was achieved without relying on extreme weights. The weight distribution was well-behaved, with a median of 0.971, interquartile range of $[0.91, 1.04]$, and only 1.5% of weights exceeding 2.0.

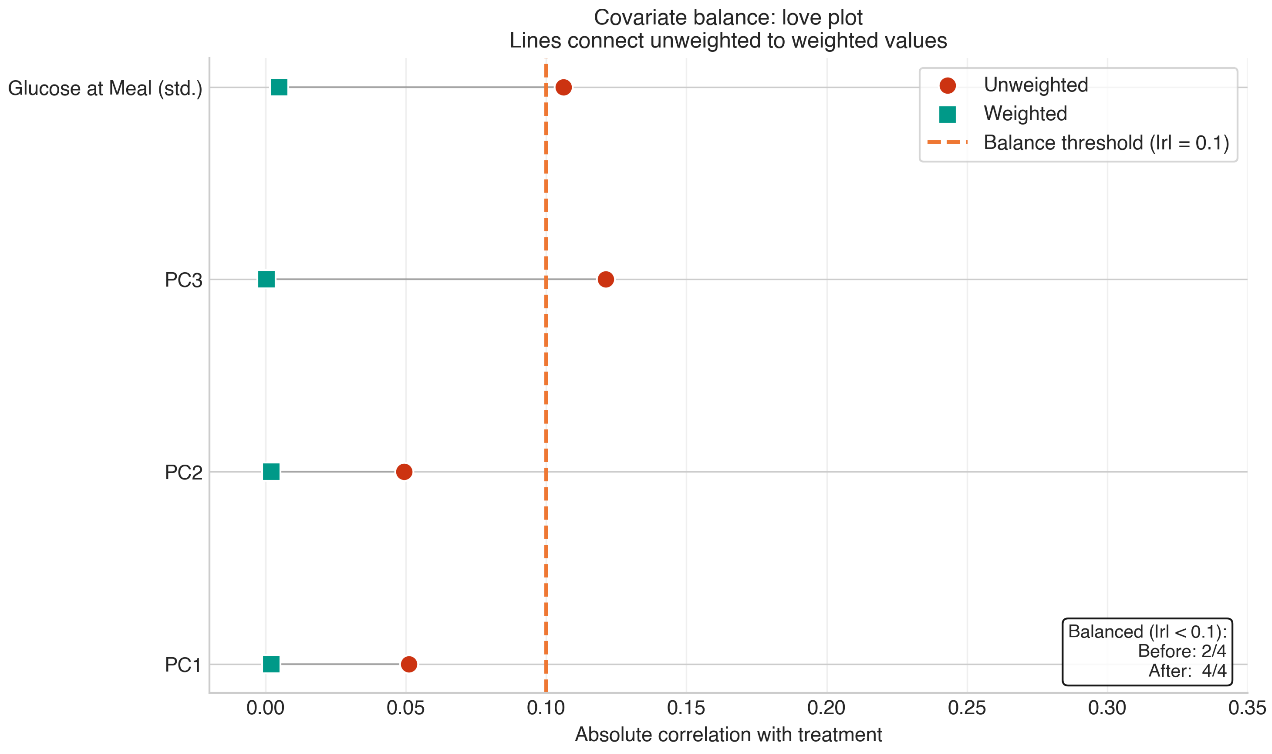


**Supplementary Figure 2: Love plot of covariate balance before and after npCBPS weighting (PCs 1–3).**

**Supplementary Table 2:** npCBPS covariate balance before and after weighting.

| Covariate | $r$ (Unwtd) | $r$ (Wtd) | Red. (%) | Status |
|---|---|---|---|---|
| PC1 | $-0.051$ | 0.002 | 96.0 | Balanced |
| PC2 | $-0.049$ | $-0.002$ | 95.9 | Balanced |
| PC3 | $-0.121$ | 0.000 | 99.7 | Balanced |
| Glucose at Meal (std.) | 0.106 | 0.005 | 95.4 | Balanced |

*Note:* $N = 199$, ESS $= 190.1$ (95.5%), weight mean (SD) $= 1.000$ (0.217), weight range $= [0.448,\ 2.093]$. Balance achieved when $|r| < 0.1$. ESS $= (\sum w_i)^2 / \sum w_i^2$.

### C Autoencoder Embedding Quality

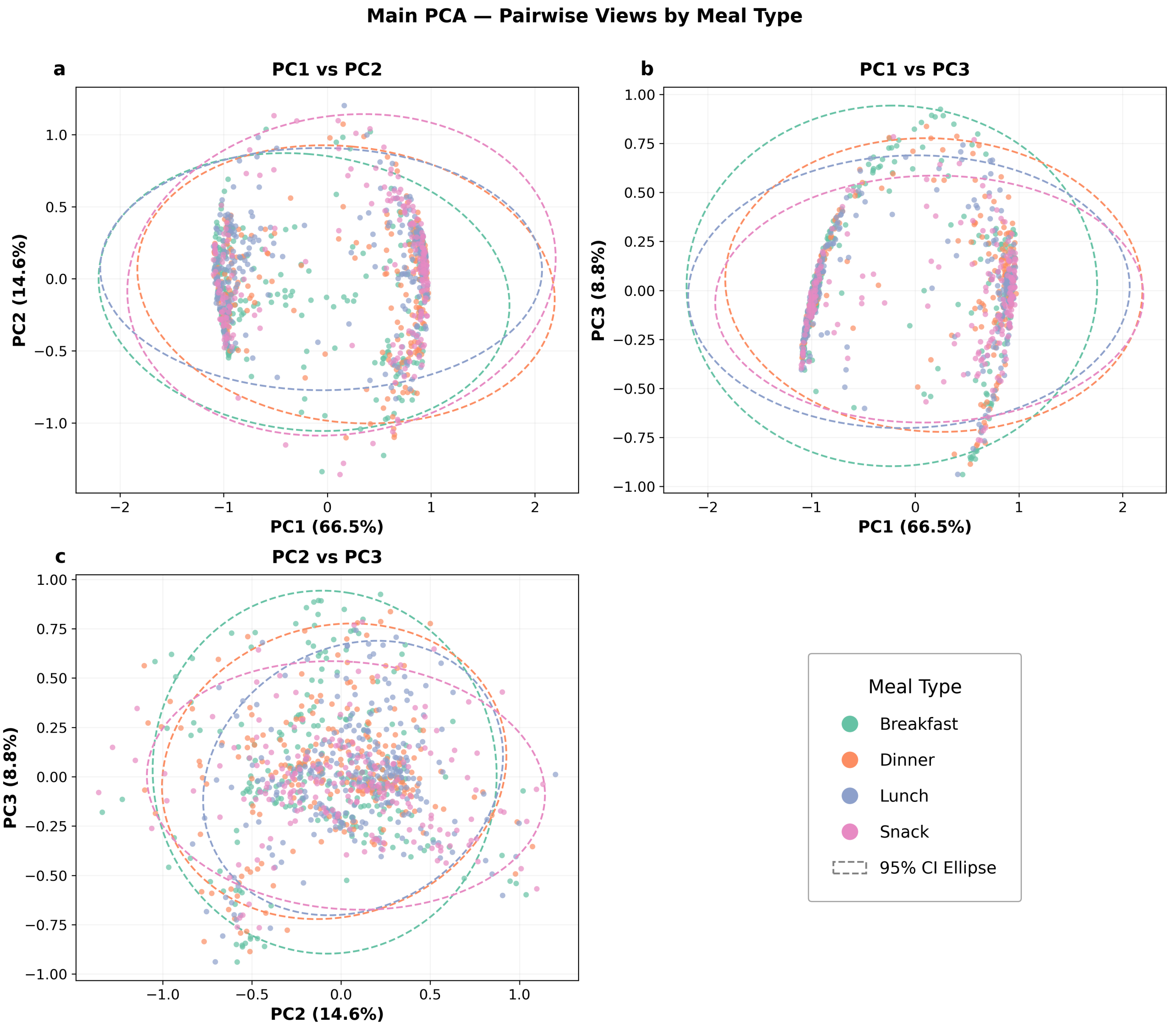


**Supplementary Figure 3: Pairwise PCA projections of learned autoencoder embeddings $\varphi$ colored by meal type (test set, $N = 199$).** Left: PC1 vs. PC2; Center: PC1 vs. PC3; Right: PC2 vs. PC3. Ellipses represent approximate 95% concentration regions. The first three principal components capture 89.9% of the total latent variance.

### D Causal Mediation Analysis Results

All models use PCA-based covariates (3 principal components capturing 90% of autoencoder latent variance) with npCBPS inverse probability weights for confounding adjustment.

**Notation and conventions.** Time = postprandial measurement time point in minutes after meal start. Dose = hypothetical increase in carbohydrate intake (grams) above the meal-type-specific median. Quantile = conditional quantile ($\tau$) of the glucose response distribution being modeled. ACME = Average Causal Mediation Effect (indirect effect mediated through insulin); ADE = Average Direct Effect (effect not mediated through insulin); Total = ACME + ADE. Est. = point estimate (mg/dL); 95% CI from quasi-Bayesian approximation with 1,000 Monte Carlo simulations; $p$ = two-sided $p$-value testing the null hypothesis that the effect equals zero. Significant results ($p < 0.05$) are shown in **bold**. The mediator model is a Tobit regression (`survreg`, Gaussian) to account for the left-censored insulin bolus distribution ($\sim$12% zeros). The outcome model is a linear mixed-effects model (LMER) with a subject-level random intercept for mean-effect results and quantile regression for distributional results. In all figures, star markers (★) denote significance ($p < 0.05$); Green = ACME; Red = ADE; Blue = Total effect. The LMER tables report all three treatment contrasts (+15, +30, +45 g). The quantile regression tables report the +30 g contrast across $\tau \in \{0.25, 0.50, 0.75\}$; full results at +15 g and +45 g contrasts are available upon request.

#### D.1 Pooled (All Meal Types)

##### D.1.1 Mean Effects (LMER Outcome)

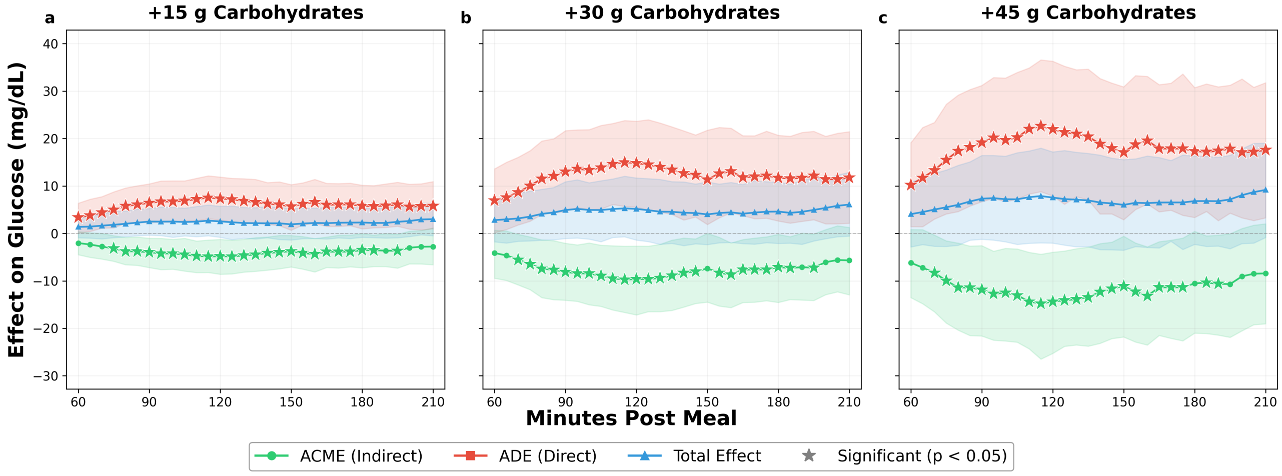


**Supplementary Figure 4: Mean causal mediation effects — Pooled (All Meal Types).** Panels show treatment contrasts (+15, +30, +45 g).

**Supplementary Table 3: LMER causal mediation effects — Pooled (all meals) ($N = 190$ meal observations, all contrast sizes).** ACME = Average Causal Mediation Effect; ADE = Average Direct Effect; Total = ACME + ADE. Est. = point estimate (mg/dL); CI = confidence interval. Significant results ($p < 0.05$) are shown in bold.

| | | ACME | | | ADE | | | Total | | |
|---|---|---|---|---|---|---|---|---|---|---|
| Time | Dose | Est. | 95% CI | $p$ | Est. | 95% CI | $p$ | Est. | 95% CI | $p$ |
| 60 min | $+15\,g$ | -2.03 | (-4.49, 0.39) | 0.10 | **3.41** | **(0.08, 6.38)** | **0.04** | 1.38 | (-0.92, 3.54) | 0.22 |
| | $+30\,g$ | -4.13 | (-9.44, 0.67) | 0.10 | **6.94** | **(0.16, 13.61)** | **0.04** | 2.81 | (-1.77, 7.31) | 0.25 |
| | $+45\,g$ | -6.17 | (-13.52, 0.83) | 0.10 | **10.24** | **(1.44, 19.11)** | **0.03** | 4.07 | (-2.85, 10.87) | 0.25 |
| 90 min | $+15\,g$ | **-3.95** | **(-7.19, -1.01)** | **0.01** | **6.45** | **(2.29, 10.44)** | **<0.001** | 2.51 | (-0.37, 5.25) | 0.08 |
| | $+30\,g$ | **-8.10** | **(-14.08, -2.08)** | **0.01** | **13.04** | **(4.67, 21.66)** | **<0.001** | 4.94 | (-1.06, 10.90) | 0.11 |
| | $+45\,g$ | **-11.85** | **(-21.61, -2.57)** | **0.01** | **19.17** | **(6.69, 31.19)** | **0.002** | 7.32 | (-1.22, 16.44) | 0.10 |
| 120 min | $+15\,g$ | **-4.81** | **(-8.60, -1.26)** | **0.01** | **7.35** | **(2.84, 11.89)** | **0.002** | 2.54 | (-0.81, 5.72) | 0.13 |
| | $+30\,g$ | **-9.64** | **(-17.17, -2.67)** | **0.01** | **14.81** | **(5.55, 23.65)** | **<0.001** | 5.17 | (-1.41, 11.69) | 0.13 |
| | $+45\,g$ | **-14.38** | **(-25.28, -3.96)** | **0.008** | **21.94** | **(7.91, 36.30)** | **0.004** | 7.56 | (-2.04, 17.19) | 0.13 |
| 150 min | $+15\,g$ | **-3.75** | **(-7.07, -0.43)** | **0.04** | **5.67** | **(0.68, 10.28)** | **0.02** | 1.92 | (-1.38, 5.23) | 0.26 |
| | $+30\,g$ | -7.37 | (-14.47, 0.01) | 0.05 | **11.39** | **(1.88, 21.76)** | **0.02** | 4.01 | (-2.35, 10.87) | 0.21 |
| | $+45\,g$ | **-11.10** | **(-21.82, -0.59)** | **0.04** | **17.11** | **(2.89, 30.89)** | **0.02** | 6.01 | (-3.54, 15.56) | 0.22 |
| 180 min | $+15\,g$ | **-3.52** | **(-7.23, -0.04)** | **0.05** | **5.82** | **(0.98, 10.16)** | **0.01** | 2.30 | (-0.81, 5.43) | 0.14 |
| | $+30\,g$ | **-7.11** | **(-14.54, -0.20)** | **0.04** | **11.72** | **(2.62, 20.68)** | **0.02** | 4.61 | (-2.10, 10.82) | 0.15 |
| | $+45\,g$ | -10.53 | (-21.01, 0.44) | 0.06 | **17.35** | **(3.37, 30.73)** | **0.02** | 6.82 | (-2.30, 16.32) | 0.16 |
| 210 min | $+15\,g$ | -2.77 | (-6.59, 1.15) | 0.15 | **5.79** | **(1.00, 10.93)** | **0.02** | 3.01 | (-0.48, 6.29) | 0.09 |
| | $+30\,g$ | -5.68 | (-12.93, 1.26) | 0.10 | **11.77** | **(2.15, 21.45)** | **0.03** | 6.09 | (-0.56, 12.88) | 0.07 |
| | $+45\,g$ | -8.42 | (-19.05, 2.03) | 0.11 | **17.59** | **(3.30, 31.76)** | **0.02** | 9.17 | (-0.85, 19.02) | 0.08 |

##### D.1.2 Quantile Regression Effects

**Supplementary Table 4: QR causal mediation effects — Pooled (all meals) ($N = 190$ meal observations, $+30\,\mathrm{g}$ contrast).** ACME = Average Causal Mediation Effect; ADE = Average Direct Effect; Total = ACME + ADE. Est. = point estimate (mg/dL); CI = confidence interval. Significant results ($p < 0.05$) are shown in bold.

| | | ACME | | | ADE | | | Total | | |
|---|---|---|---|---|---|---|---|---|---|---|
| Time | Quantile | Est. | 95% CI | $p$ | Est. | 95% CI | $p$ | Est. | 95% CI | $p$ |
| 60 min | $\tau = 0.25$ | -2.65 | (-10.45, 5.09) | 0.50 | 6.84 | (-2.62, 16.26) | 0.16 | 4.20 | (-1.17, 9.75) | 0.11 |
| | $\tau = 0.50$ | -0.38 | (-7.28, 7.15) | 0.88 | 5.66 | (-4.38, 15.52) | 0.26 | 5.28 | (-0.98, 11.55) | 0.09 |
| | $\tau = 0.75$ | **-9.30** | **(-13.24, -5.79)** | **<0.001** | **12.93** | **(9.67, 16.42)** | **<0.001** | **3.63** | **(0.97, 6.17)** | **0.01** |
| 90 min | $\tau = 0.25$ | -7.33 | (-17.65, 2.36) | 0.15 | 12.17 | (-3.44, 28.29) | 0.14 | 4.84 | (-6.40, 16.09) | 0.43 |
| | $\tau = 0.50$ | **-9.46** | **(-18.00, -0.91)** | **0.04** | **13.83** | **(3.12, 23.97)** | **0.01** | 4.36 | (-1.19, 9.57) | 0.11 |
| | $\tau = 0.75$ | **-10.81** | **(-16.24, -5.69)** | **<0.001** | **19.33** | **(9.02, 29.75)** | **<0.001** | 8.52 | (-0.06, 17.82) | 0.05 |
| 120 min | $\tau = 0.25$ | -7.34 | (-17.41, 2.75) | 0.13 | 10.95 | (-3.10, 25.01) | 0.15 | 3.61 | (-6.89, 13.65) | 0.52 |
| | $\tau = 0.50$ | **-10.06** | **(-15.99, -4.36)** | **<0.001** | **19.71** | **(9.24, 30.03)** | **<0.001** | **9.66** | **(0.68, 18.36)** | **0.04** |
| | $\tau = 0.75$ | -5.79 | (-17.21, 5.98) | 0.31 | **19.73** | **(4.43, 33.85)** | **0.02** | **13.95** | **(4.82, 23.47)** | **0.006** |
| 150 min | $\tau = 0.25$ | -8.19 | (-20.17, 3.92) | 0.19 | 8.93 | (-6.89, 23.68) | 0.28 | 0.74 | (-5.95, 7.44) | 0.85 |
| | $\tau = 0.50$ | -6.06 | (-12.34, 0.06) | 0.05 | 9.20 | (-1.69, 20.24) | 0.10 | 3.14 | (-6.59, 12.74) | 0.50 |
| | $\tau = 0.75$ | -7.79 | (-17.77, 2.15) | 0.12 | **18.09** | **(5.07, 31.07)** | **0.006** | **10.29** | **(2.14, 18.90)** | **0.02** |
| 180 min | $\tau = 0.25$ | **-10.08** | **(-18.43, -2.52)** | **0.008** | **14.69** | **(2.26, 27.18)** | **0.02** | 4.62 | (-4.70, 13.08) | 0.28 |
| | $\tau = 0.50$ | -2.62 | (-12.88, 6.87) | 0.61 | 5.85 | (-8.56, 20.02) | 0.43 | 3.22 | (-5.82, 12.35) | 0.48 |
| | $\tau = 0.75$ | -5.00 | (-12.49, 2.03) | 0.17 | **12.82** | **(3.75, 21.39)** | **0.004** | **7.82** | **(2.58, 13.31)** | **0.002** |
| 210 min | $\tau = 0.25$ | -9.53 | (-20.58, 0.78) | 0.08 | 15.50 | (-0.63, 31.36) | 0.07 | 5.97 | (-4.23, 16.25) | 0.26 |
| | $\tau = 0.50$ | -6.73 | (-14.95, 1.70) | 0.10 | **14.44** | **(2.95, 26.07)** | **0.01** | 7.71 | (-0.18, 15.70) | 0.06 |
| | $\tau = 0.75$ | -2.33 | (-7.84, 2.99) | 0.41 | **15.12** | **(8.20, 21.95)** | **<0.001** | **12.79** | **(5.77, 19.41)** | **<0.001** |

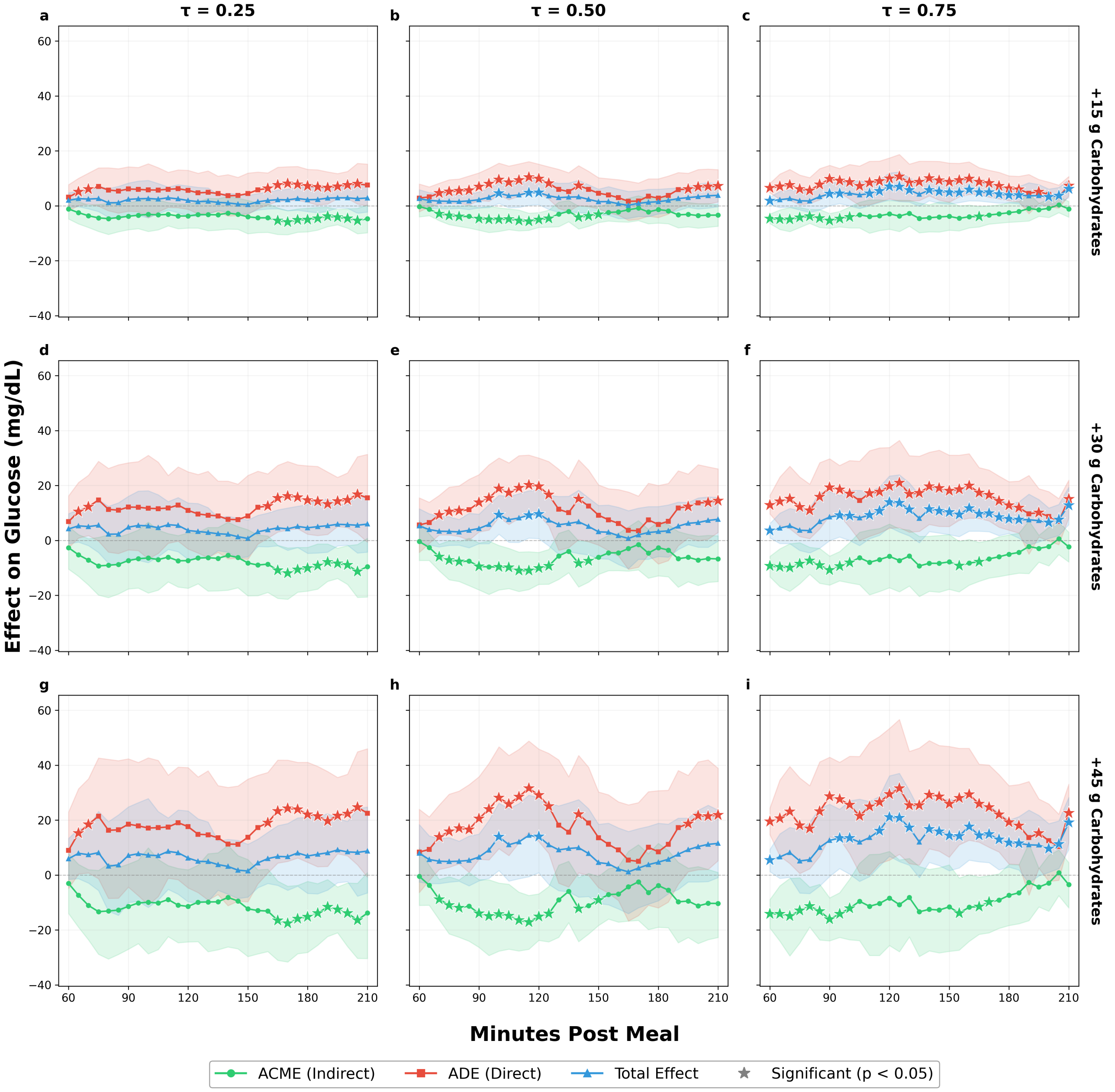


**Supplementary Figure 5: Quantile regression mediation effects — Pooled (All Meal Types).** Rows: treatment contrasts (+15, +30, +45 g). Columns: $\tau \in \{0.25, 0.50, 0.75\}$.

#### D.2 Dinner

##### D.2.1 Mean Effects (LMER Outcome)

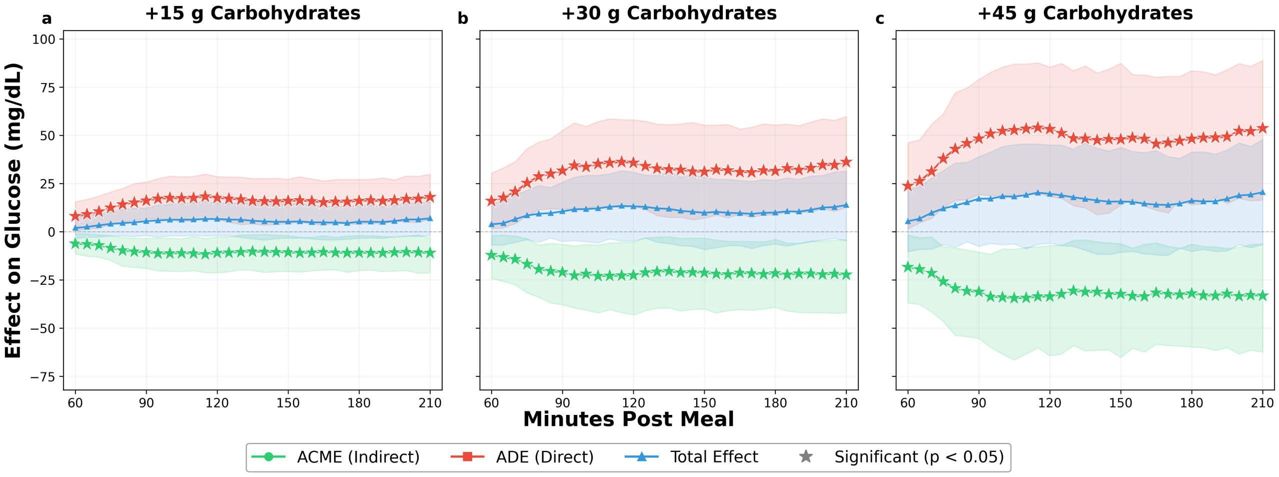


**Supplementary Figure 6: Mean causal mediation effects — Dinner.** Panels show treatment contrasts (+15, +30, +45 g).

**Supplementary Table 5: LMER causal mediation effects — Dinner ($N = 42$ meal observations, all contrast sizes).** ACME = Average Causal Mediation Effect; ADE = Average Direct Effect; Total = ACME + ADE. Est. = point estimate (mg/dL); CI = confidence interval. Significant results ($p < 0.05$) are shown in bold.

| | | ACME | | | ADE | | | Total | | |
|---|---|---|---|---|---|---|---|---|---|---|
| Time | Dose | Est. | 95% CI | $p$ | Est. | 95% CI | $p$ | Est. | 95% CI | $p$ |
| 60 min | +15 $g$ | **-6.15** | **(-11.53, -1.00)** | **0.02** | **8.14** | **(0.51, 15.63)** | **0.03** | 1.99 | (-3.41, 7.56) | 0.48 |
| | +30 $g$ | **-12.09** | **(-24.18, -1.95)** | **0.02** | **15.96** | **(1.69, 30.46)** | **0.03** | 3.87 | (-6.81, 14.75) | 0.45 |
| | +45 $g$ | **-18.32** | **(-36.95, -1.65)** | **0.03** | **23.75** | **(1.49, 46.17)** | **0.05** | 5.43 | (-10.32, 22.21) | 0.50 |
| 90 min | +15 $g$ | **-10.64** | **(-18.99, -3.35)** | **0.002** | **16.17** | **(6.91, 25.84)** | **<0.001** | 5.53 | (-1.91, 12.36) | 0.15 |
| | +30 $g$ | **-21.07** | **(-37.83, -6.52)** | **0.01** | **31.64** | **(11.73, 52.61)** | **0.002** | 10.57 | (-5.05, 25.68) | 0.18 |
| | +45 $g$ | **-31.23** | **(-55.36, -10.48)** | **0.004** | **48.39** | **(19.43, 79.05)** | **0.002** | 17.15 | (-7.55, 38.99) | 0.15 |
| 120 min | +15 $g$ | **-10.96** | **(-21.32, -2.56)** | **0.008** | **17.58** | **(6.54, 28.61)** | **0.004** | 6.61 | (-1.87, 14.93) | 0.11 |
| | +30 $g$ | **-22.56** | **(-43.14, -5.43)** | **0.01** | **35.73** | **(13.42, 58.06)** | **0.006** | 13.17 | (-4.77, 30.87) | 0.14 |
| | +45 $g$ | **-33.65** | **(-64.29, -7.12)** | **0.008** | **53.34** | **(18.22, 85.44)** | **0.004** | 19.69 | (-5.88, 45.00) | 0.14 |
| 150 min | +15 $g$ | **-10.73** | **(-20.55, -2.04)** | **0.02** | **15.94** | **(4.36, 27.29)** | **0.006** | 5.21 | (-3.28, 14.49) | 0.23 |
| | +30 $g$ | **-21.26** | **(-40.08, -3.30)** | **0.02** | **31.11** | **(7.09, 55.33)** | **0.02** | 9.84 | (-9.19, 28.31) | 0.30 |
| | +45 $g$ | **-32.26** | **(-65.22, -6.18)** | **0.01** | **47.98** | **(13.94, 87.38)** | **<0.001** | 15.72 | (-10.65, 43.71) | 0.25 |
| 180 min | +15 $g$ | **-10.84** | **(-20.16, -1.89)** | **0.02** | **16.04** | **(4.09, 27.16)** | **0.006** | 5.21 | (-3.39, 13.40) | 0.23 |
| | +30 $g$ | **-21.53** | **(-39.27, -3.94)** | **0.02** | **31.49** | **(8.76, 55.41)** | **0.004** | 9.96 | (-6.36, 26.66) | 0.25 |
| | +45 $g$ | **-32.04** | **(-59.83, -6.42)** | **0.02** | **48.20** | **(13.98, 83.55)** | **0.008** | 16.16 | (-10.26, 41.32) | 0.23 |
| 210 min | +15 $g$ | **-11.05** | **(-21.33, -3.15)** | **0.008** | **17.99** | **(6.74, 29.84)** | **<0.001** | 6.94 | (-2.01, 15.35) | 0.13 |
| | +30 $g$ | **-22.30** | **(-42.02, -4.14)** | **0.01** | **36.20** | **(12.67, 59.92)** | **0.002** | 13.90 | (-4.60, 31.52) | 0.14 |
| | +45 $g$ | **-33.20** | **(-62.42, -6.24)** | **0.02** | **53.70** | **(16.47, 88.88)** | **0.008** | 20.50 | (-6.60, 47.88) | 0.14 |

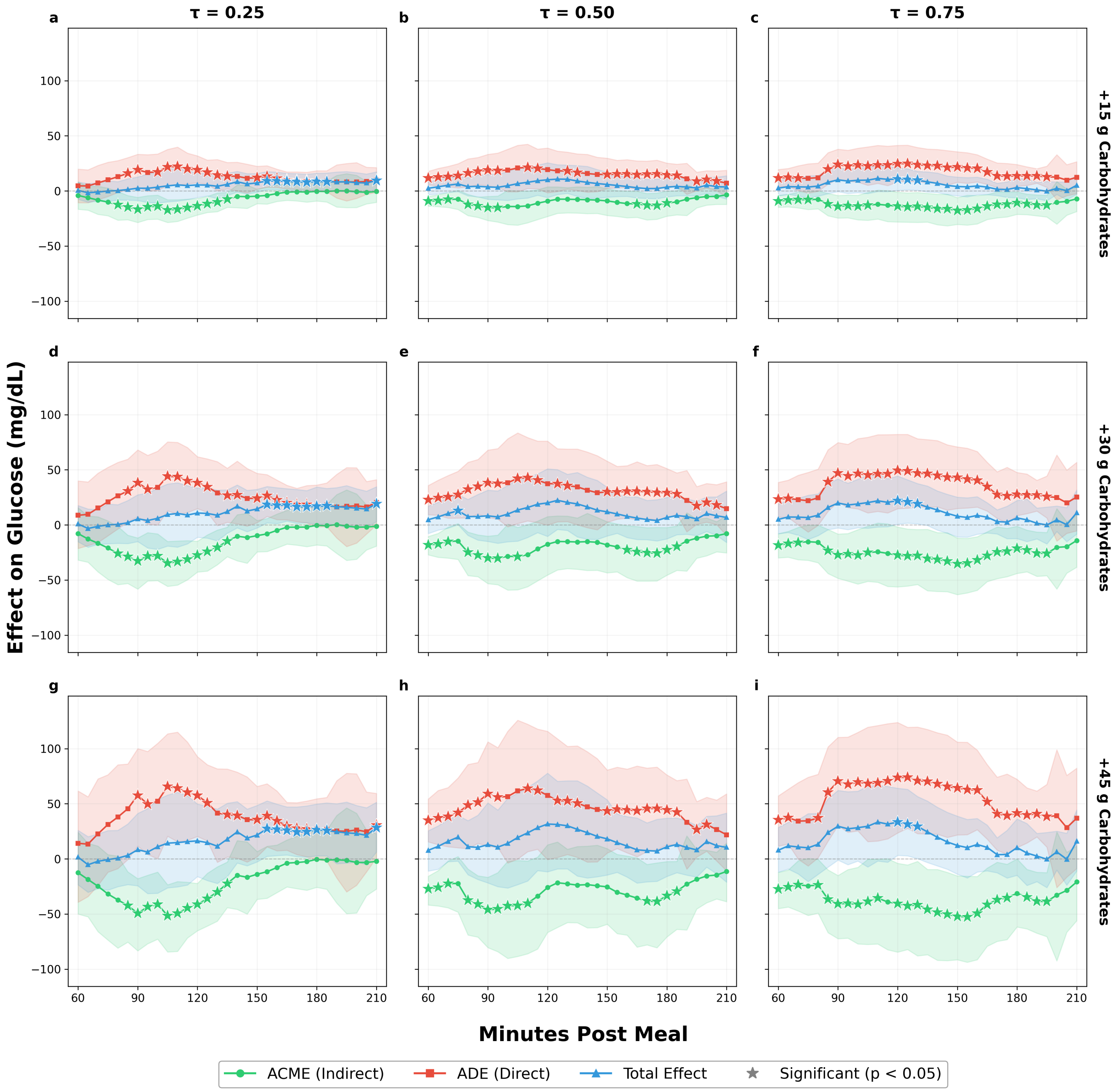


**Supplementary Figure 7: Quantile regression mediation effects — Dinner.** Rows: treatment contrasts (+15, +30, +45 g). Columns: $\tau \in \{0.25, 0.50, 0.75\}$.

##### D.2.2 Quantile Regression Effects

**Supplementary Table 6: QR causal mediation effects — Dinner ($N = 42$ meal observations, $+30$ g contrast).** ACME = Average Causal Mediation Effect; ADE = Average Direct Effect; Total = ACME + ADE. Est. = point estimate (mg/dL); CI = confidence interval. Significant results ($p < 0.05$) are shown in bold.

| | | ACME | | | ADE | | | Total | | |
|---|---|---|---|---|---|---|---|---|---|---|
| Time | Quantile | Est. | 95% CI | $p$ | Est. | 95% CI | $p$ | Est. | 95% CI | $p$ |
| 60 min | $\tau = 0.25$ | -7.86 | (-31.91, 15.55) | 0.50 | 8.70 | (-21.36, 39.89) | 0.61 | 0.84 | (-15.26, 17.59) | 0.93 |
| | $\tau = 0.50$ | **-18.12** | **(-27.15, -10.52)** | **<0.001** | **22.82** | **(9.24, 35.70)** | **<0.001** | 4.70 | (-7.89, 17.05) | 0.45 |
| | $\tau = 0.75$ | **-18.37** | **(-29.94, -7.95)** | **<0.001** | **23.39** | **(7.11, 38.56)** | **0.004** | 5.02 | (-8.10, 18.96) | 0.45 |
| 90 min | $\tau = 0.25$ | **-32.78** | **(-58.22, -11.84)** | **0.002** | **38.30** | **(7.65, 67.99)** | **0.01** | 5.52 | (-15.12, 27.33) | 0.61 |
| | $\tau = 0.50$ | **-30.07** | **(-53.28, -10.24)** | **<0.001** | **38.28** | **(6.73, 67.83)** | **0.02** | 8.21 | (-14.99, 31.48) | 0.47 |
| | $\tau = 0.75$ | **-27.16** | **(-48.46, -8.29)** | **0.004** | **46.98** | **(20.79, 74.66)** | **<0.001** | 19.81 | (-2.78, 41.60) | 0.07 |
| 120 min | $\tau = 0.25$ | **-27.49** | **(-42.78, -14.81)** | **<0.001** | **38.28** | **(14.79, 61.82)** | **0.004** | 10.79 | (-11.23, 33.41) | 0.35 |
| | $\tau = 0.50$ | -17.49 | (-45.49, 7.49) | 0.18 | 37.44 | (-0.23, 76.00) | 0.06 | 19.95 | (-10.44, 50.84) | 0.17 |
| | $\tau = 0.75$ | **-27.43** | **(-56.25, -1.94)** | **0.04** | **49.46** | **(14.69, 82.08)** | **0.008** | **22.03** | **(0.59, 44.31)** | **0.04** |
| 150 min | $\tau = 0.25$ | -9.75 | (-24.50, 4.03) | 0.18 | **24.20** | **(2.00, 46.73)** | **0.03** | 14.46 | (-2.78, 32.24) | 0.11 |
| | $\tau = 0.50$ | -18.20 | (-43.49, 2.84) | 0.10 | **30.02** | **(2.31, 57.77)** | **0.04** | 11.82 | (-10.06, 34.02) | 0.34 |
| | $\tau = 0.75$ | **-35.24** | **(-63.25, -12.01)** | **0.002** | **43.09** | **(14.87, 70.75)** | **0.004** | 7.85 | (-10.87, 25.14) | 0.38 |
| 180 min | $\tau = 0.25$ | -0.38 | (-18.20, 16.98) | 0.94 | 17.24 | (-1.76, 36.14) | 0.09 | **16.87** | **(0.52, 34.38)** | **0.05** |
| | $\tau = 0.50$ | **-22.59** | **(-45.66, -2.41)** | **0.03** | **29.31** | **(6.19, 51.10)** | **0.02** | 6.72 | (-12.69, 24.93) | 0.46 |
| | $\tau = 0.75$ | **-21.27** | **(-44.04, -1.24)** | **0.03** | **27.59** | **(8.42, 47.55)** | **0.008** | 6.32 | (-12.42, 22.17) | 0.46 |
| 210 min | $\tau = 0.25$ | -1.30 | (-19.24, 16.20) | 0.89 | 20.38 | (-0.06, 41.00) | 0.06 | **19.09** | **(2.99, 34.61)** | **0.01** |
| | $\tau = 0.50$ | -7.85 | (-25.38, 8.65) | 0.34 | 14.67 | (-9.80, 39.06) | 0.24 | 6.83 | (-15.96, 30.70) | 0.57 |
| | $\tau = 0.75$ | -14.16 | (-38.36, 8.80) | 0.20 | 25.21 | (-5.42, 56.73) | 0.13 | 11.05 | (-8.33, 31.60) | 0.27 |

#### D.3 Breakfast

##### D.3.1 Mean Effects (LMER Outcome)

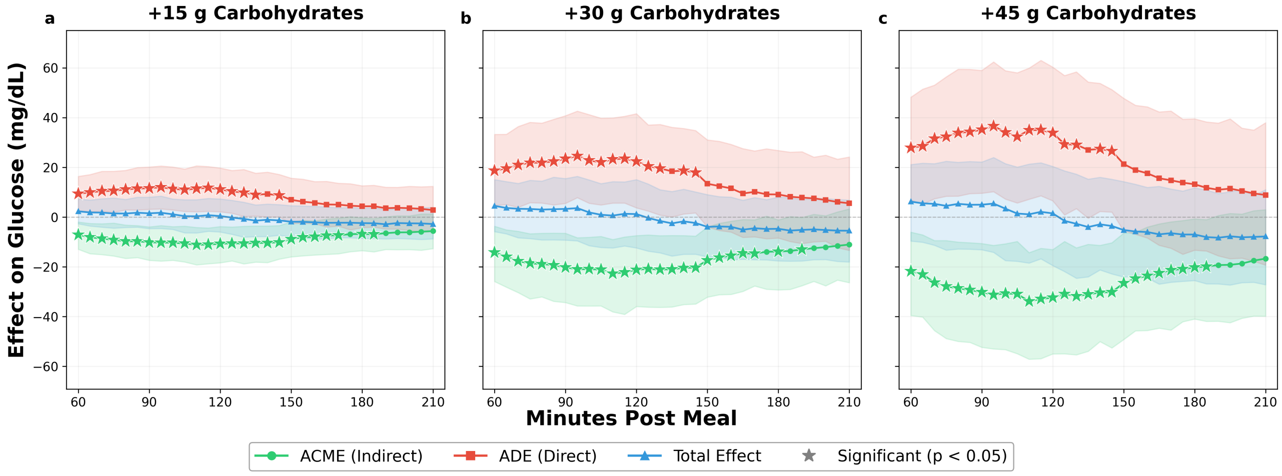


**Supplementary Figure 8: Mean causal mediation effects — Breakfast.** Panels show treatment contrasts ($+15$, $+30$, $+45$ g).

**Supplementary Table 7: LMER causal mediation effects — Breakfast ($N = 51$ meal observations, all contrast sizes).** ACME = Average Causal Mediation Effect; ADE = Average Direct Effect; Total = ACME + ADE. Est. = point estimate (mg/dL); CI = confidence interval. Significant results ($p < 0.05$) are shown in bold.

| | | ACME | | | ADE | | | Total | | |
|---|---|---|---|---|---|---|---|---|---|---|
| Time | Dose | Est. | 95% CI | $p$ | Est. | 95% CI | $p$ | Est. | 95% CI | $p$ |
| 60 min | $+15\,g$ | **-7.08** | **(-12.93, -1.96)** | **<0.001** | **9.40** | **(2.56, 16.39)** | **0.01** | 2.33 | (-2.67, 7.32) | 0.36 |
| | $+30\,g$ | **-14.19** | **(-25.88, -3.66)** | **0.004** | **18.72** | **(4.75, 33.26)** | **0.01** | 4.54 | (-5.83, 15.08) | 0.35 |
| | $+45\,g$ | **-21.68** | **(-39.53, -6.03)** | **0.004** | **27.92** | **(6.86, 48.22)** | **0.01** | 6.24 | (-9.62, 21.14) | 0.42 |
| 90 min | $+15\,g$ | **-10.12** | **(-17.36, -3.93)** | **0.002** | **11.65** | **(3.15, 20.15)** | **0.008** | 1.53 | (-4.61, 8.03) | 0.64 |
| | $+30\,g$ | **-20.16** | **(-34.34, -6.50)** | **0.004** | **23.46** | **(5.53, 40.78)** | **0.01** | 3.30 | (-9.24, 15.51) | 0.65 |
| | $+45\,g$ | **-30.16** | **(-52.34, -10.61)** | **0.002** | **35.15** | **(9.60, 58.98)** | **0.008** | 5.00 | (-13.12, 22.17) | 0.55 |
| 120 min | $+15\,g$ | **-10.71** | **(-18.37, -3.79)** | **<0.001** | **11.13** | **(2.14, 19.74)** | **0.01** | 0.42 | (-5.97, 7.31) | 0.91 |
| | $+30\,g$ | **-21.15** | **(-35.93, -7.43)** | **0.004** | **22.38** | **(3.72, 41.58)** | **0.02** | 1.23 | (-12.39, 15.09) | 0.85 |
| | $+45\,g$ | **-32.34** | **(-55.01, -13.04)** | **0.002** | **33.85** | **(6.56, 60.33)** | **0.01** | 1.51 | (-19.47, 20.40) | 0.88 |
| 150 min | $+15\,g$ | **-8.81** | **(-16.00, -1.90)** | **0.02** | 6.96 | (-2.58, 15.66) | 0.13 | -1.85 | (-8.06, 4.60) | 0.55 |
| | $+30\,g$ | **-17.34** | **(-32.17, -3.95)** | **0.006** | 13.43 | (-3.97, 31.04) | 0.12 | -3.91 | (-15.96, 9.05) | 0.54 |
| | $+45\,g$ | **-26.55** | **(-49.23, -6.41)** | **0.004** | 21.36 | (-5.53, 47.79) | 0.13 | -5.19 | (-24.07, 14.28) | 0.59 |
| 180 min | $+15\,g$ | **-6.76** | **(-13.48, -0.29)** | **0.04** | 4.36 | (-4.23, 12.60) | 0.32 | -2.40 | (-8.56, 3.51) | 0.44 |
| | $+30\,g$ | **-13.82** | **(-29.26, -1.13)** | **0.03** | 9.12 | (-8.40, 26.78) | 0.30 | -4.70 | (-17.61, 8.04) | 0.45 |
| | $+45\,g$ | **-20.20** | **(-40.97, -0.69)** | **0.04** | 13.21 | (-13.28, 39.53) | 0.32 | -7.00 | (-25.50, 11.77) | 0.47 |
| 210 min | $+15\,g$ | -5.60 | (-12.66, 1.27) | 0.11 | 2.83 | (-5.88, 12.37) | 0.54 | -2.77 | (-8.73, 3.93) | 0.35 |
| | $+30\,g$ | -11.03 | (-26.35, 3.30) | 0.13 | 5.56 | (-13.38, 24.19) | 0.54 | -5.47 | (-18.04, 6.86) | 0.37 |
| | $+45\,g$ | -16.68 | (-39.82, 2.99) | 0.10 | 8.90 | (-19.31, 37.97) | 0.53 | -7.78 | (-27.18, 11.07) | 0.47 |

##### D.3.2 Quantile Regression Effects

**Supplementary Table 8: QR causal mediation effects — Breakfast ($N = 51$ meal observations, $+30\,g$ contrast).** ACME = Average Causal Mediation Effect; ADE = Average Direct Effect; Total = ACME + ADE. Est. = point estimate (mg/dL); CI = confidence interval. Significant results ($p < 0.05$) are shown in bold.

| | | ACME | | | ADE | | | Total | | |
|---|---|---|---|---|---|---|---|---|---|---|
| Time | Quantile | Est. | 95% CI | $p$ | Est. | 95% CI | $p$ | Est. | 95% CI | $p$ |
| 60 min | $\tau = 0.25$ | -14.93 | (-33.50, 3.87) | 0.11 | **29.09** | **(7.02, 49.27)** | **0.01** | **14.16** | **(2.79, 25.87)** | **0.01** |
| | $\tau = 0.50$ | -11.99 | (-30.61, 6.83) | 0.18 | 16.90 | (-5.37, 38.09) | 0.13 | 4.91 | (-3.69, 13.86) | 0.28 |
| | $\tau = 0.75$ | -11.55 | (-25.62, 1.90) | 0.08 | **15.11** | **(1.02, 29.49)** | **0.04** | 3.56 | (-2.78, 9.53) | 0.24 |
| 90 min | $\tau = 0.25$ | -16.45 | (-33.68, 0.14) | 0.05 | **22.64** | **(2.50, 43.72)** | **0.03** | 6.19 | (-6.26, 20.01) | 0.36 |
| | $\tau = 0.50$ | -15.77 | (-33.87, 0.51) | 0.06 | 12.68 | (-7.79, 32.92) | 0.22 | -3.09 | (-12.77, 7.02) | 0.54 |
| | $\tau = 0.75$ | **-21.34** | **(-29.72, -14.11)** | **<0.001** | **20.43** | **(9.92, 31.03)** | **<0.001** | -0.92 | (-12.56, 10.86) | 0.87 |
| 120 min | $\tau = 0.25$ | -10.28 | (-35.94, 14.86) | 0.38 | 17.53 | (-14.02, 48.19) | 0.24 | 7.25 | (-4.52, 19.27) | 0.23 |
| | $\tau = 0.50$ | **-17.21** | **(-33.81, -2.49)** | **0.02** | **16.40** | **(1.87, 30.89)** | **0.03** | -0.81 | (-9.55, 7.28) | 0.88 |
| | $\tau = 0.75$ | **-24.58** | **(-43.58, -8.31)** | **<0.001** | 23.58 | (-2.20, 49.91) | 0.09 | -1.00 | (-22.60, 21.57) | 0.96 |
| 150 min | $\tau = 0.25$ | -9.35 | (-36.19, 15.70) | 0.46 | 0.87 | (-27.30, 30.56) | 0.95 | -8.48 | (-17.96, 0.92) | 0.08 |
| | $\tau = 0.50$ | **-16.02** | **(-31.63, -3.91)** | **0.01** | 11.02 | (-1.57, 23.59) | 0.10 | -5.00 | (-16.70, 5.11) | 0.37 |
| | $\tau = 0.75$ | **-19.54** | **(-32.91, -7.22)** | **<0.001** | 15.19 | (-5.50, 36.32) | 0.15 | -4.34 | (-26.89, 18.80) | 0.72 |
| 180 min | $\tau = 0.25$ | -4.76 | (-20.64, 10.92) | 0.54 | 0.98 | (-16.54, 17.80) | 0.89 | -3.78 | (-17.74, 11.07) | 0.58 |
| | $\tau = 0.50$ | **-16.91** | **(-25.81, -9.20)** | **<0.001** | 6.52 | (-5.73, 20.45) | 0.32 | -10.40 | (-22.30, 1.98) | 0.12 |
| | $\tau = 0.75$ | -12.59 | (-30.87, 5.47) | 0.17 | 8.67 | (-16.19, 33.44) | 0.52 | -3.92 | (-22.69, 17.41) | 0.68 |
| 210 min | $\tau = 0.25$ | -5.28 | (-24.74, 11.97) | 0.59 | 5.29 | (-9.14, 20.38) | 0.48 | 0.01 | (-7.97, 6.45) | 0.95 |
| | $\tau = 0.50$ | -10.63 | (-36.60, 11.40) | 0.38 | 7.59 | (-18.49, 35.95) | 0.55 | -3.03 | (-14.75, 8.96) | 0.58 |
| | $\tau = 0.75$ | **-14.66** | **(-23.39, -6.51)** | **<0.001** | 8.32 | (-16.97, 32.87) | 0.51 | -6.34 | (-29.38, 16.23) | 0.57 |

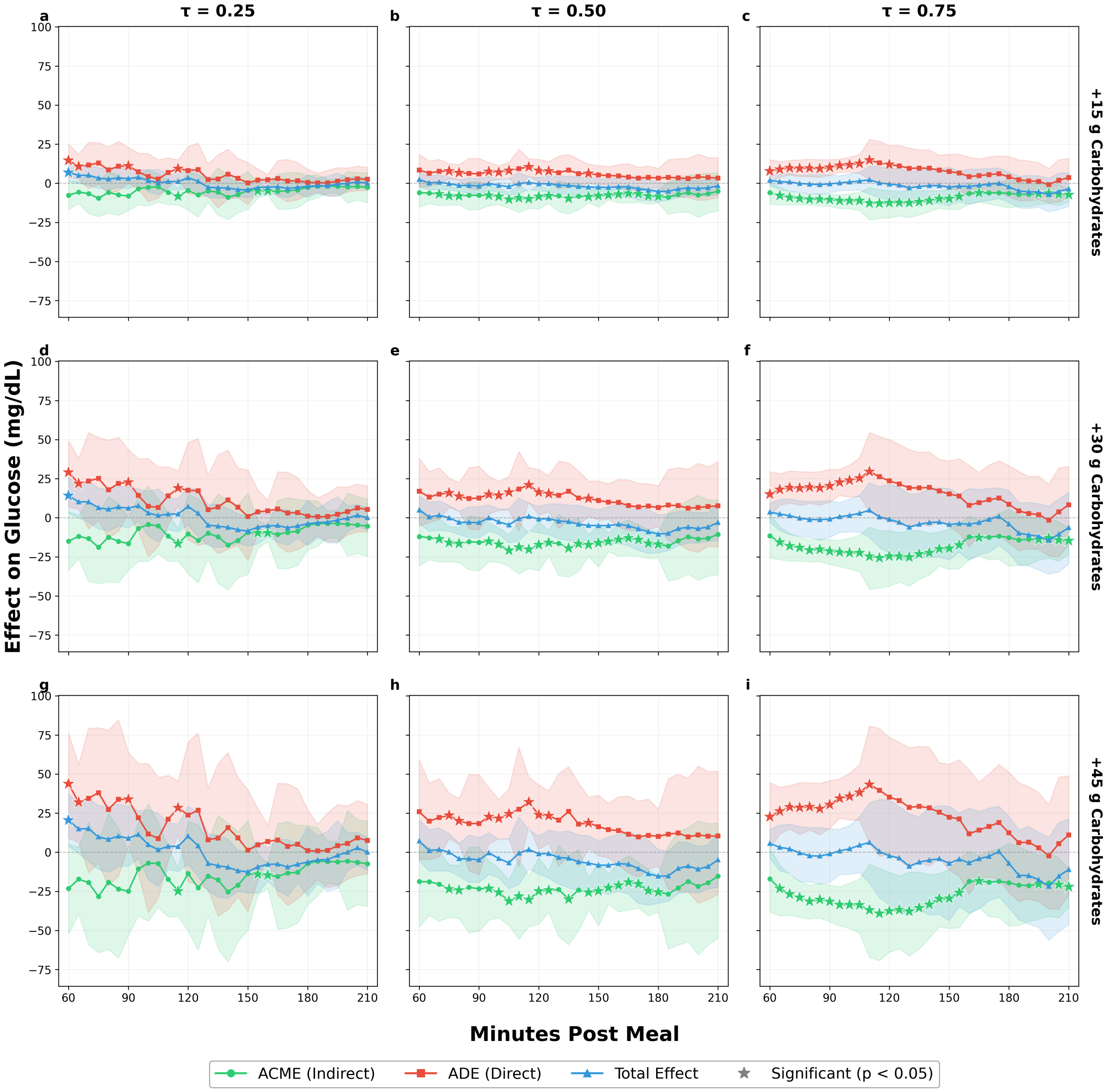


**Supplementary Figure 9: Quantile regression mediation effects — Breakfast.** Rows: treatment contrasts (+15, +30, +45 g). Columns: $\tau \in \{0.25, 0.50, 0.75\}$.

#### D.4 Lunch

##### D.4.1 Mean Effects (LMER Outcome)

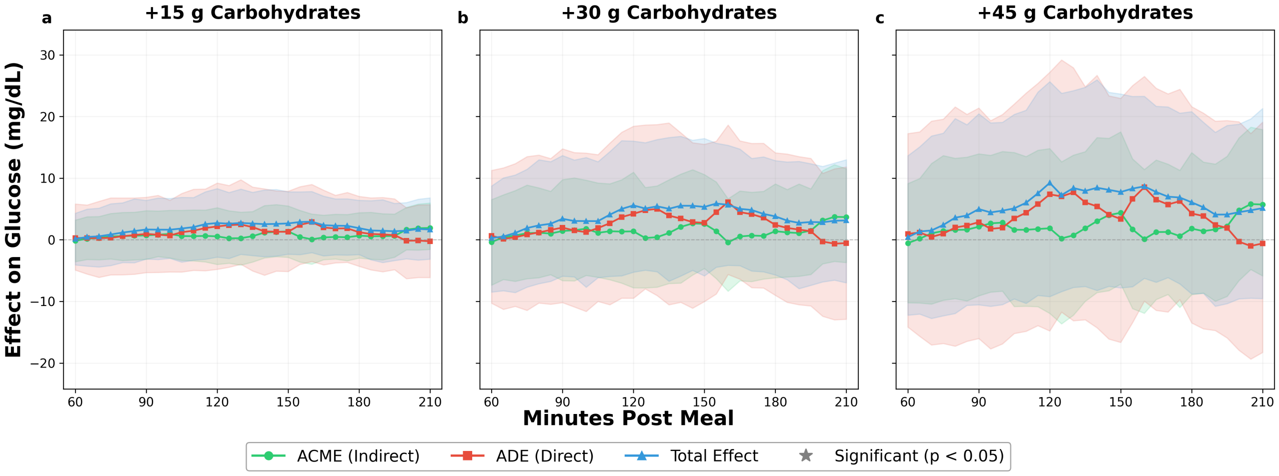


**Supplementary Figure 10: Mean causal mediation effects — Lunch.** Panels show treatment contrasts (+15, +30, +45 g).

**Supplementary Table 9: LMER causal mediation effects — Lunch ($N = 58$ meal observations, all contrast sizes).** ACME = Average Causal Mediation Effect; ADE = Average Direct Effect; Total = ACME + ADE. Est. = point estimate (mg/dL); CI = confidence interval. Significant results ($p < 0.05$) are shown in bold.

| | | ACME | | | ADE | | | Total | | |
|---|---|---|---|---|---|---|---|---|---|---|
| Time | Dose | Est. | 95% CI | $p$ | Est. | 95% CI | $p$ | Est. | 95% CI | $p$ |
| 60 min | +15 $g$ | -0.16 | (-3.55, 3.19) | 0.90 | 0.30 | (-4.92, 5.81) | 0.92 | 0.14 | (-4.08, 4.30) | 0.95 |
| | +30 $g$ | -0.37 | (-7.36, 6.52) | 0.91 | 0.65 | (-10.29, 11.26) | 0.90 | 0.28 | (-8.49, 8.73) | 0.91 |
| | +45 $g$ | -0.55 | (-10.22, 9.12) | 0.93 | 0.96 | (-14.15, 17.23) | 0.92 | 0.42 | (-12.23, 13.64) | 0.96 |
| 90 min | +15 $g$ | 0.71 | (-3.17, 4.72) | 0.73 | 0.95 | (-5.32, 6.89) | 0.76 | 1.66 | (-3.12, 6.39) | 0.49 |
| | +30 $g$ | 1.42 | (-6.63, 9.75) | 0.69 | 1.98 | (-10.19, 14.76) | 0.75 | 3.40 | (-5.91, 13.67) | 0.52 |
| | +45 $g$ | 2.12 | (-9.09, 13.94) | 0.69 | 2.86 | (-16.02, 21.38) | 0.77 | 4.98 | (-10.55, 20.44) | 0.53 |
| 120 min | +15 $g$ | 0.55 | (-4.00, 5.20) | 0.78 | 2.17 | (-4.79, 9.21) | 0.56 | 2.72 | (-2.81, 8.30) | 0.35 |
| | +30 $g$ | 1.36 | (-7.79, 10.96) | 0.77 | 4.21 | (-9.48, 18.42) | 0.57 | 5.57 | (-5.26, 16.01) | 0.30 |
| | +45 $g$ | 1.86 | (-11.95, 14.79) | 0.78 | 7.37 | (-14.79, 27.18) | 0.50 | 9.23 | (-9.22, 25.66) | 0.29 |
| 150 min | +15 $g$ | 1.36 | (-2.85, 5.48) | 0.49 | 1.28 | (-5.15, 7.88) | 0.70 | 2.65 | (-2.34, 7.79) | 0.32 |
| | +30 $g$ | 2.59 | (-4.66, 10.58) | 0.51 | 2.74 | (-10.02, 15.49) | 0.70 | 5.33 | (-5.86, 16.43) | 0.30 |
| | +45 $g$ | 4.37 | (-7.18, 17.51) | 0.46 | 3.37 | (-16.65, 22.94) | 0.70 | 7.74 | (-7.81, 23.66) | 0.33 |
| 180 min | +15 $g$ | 0.66 | (-2.97, 4.39) | 0.72 | 1.22 | (-4.30, 7.04) | 0.68 | 1.88 | (-2.40, 6.40) | 0.41 |
| | +30 $g$ | 1.39 | (-6.16, 9.06) | 0.70 | 2.40 | (-9.01, 14.11) | 0.73 | 3.79 | (-5.71, 12.84) | 0.44 |
| | +45 $g$ | 1.81 | (-8.90, 14.15) | 0.80 | 4.27 | (-13.49, 21.60) | 0.62 | 6.08 | (-8.79, 20.79) | 0.39 |
| 210 min | +15 $g$ | 1.91 | (-1.56, 5.97) | 0.30 | -0.25 | (-6.12, 5.70) | 0.91 | 1.66 | (-3.14, 6.80) | 0.51 |
| | +30 $g$ | 3.67 | (-3.70, 11.52) | 0.33 | -0.54 | (-12.89, 11.82) | 0.95 | 3.13 | (-6.98, 12.99) | 0.55 |
| | +45 $g$ | 5.72 | (-5.86, 17.87) | 0.29 | -0.60 | (-18.27, 19.09) | 0.92 | 5.12 | (-9.54, 21.30) | 0.54 |

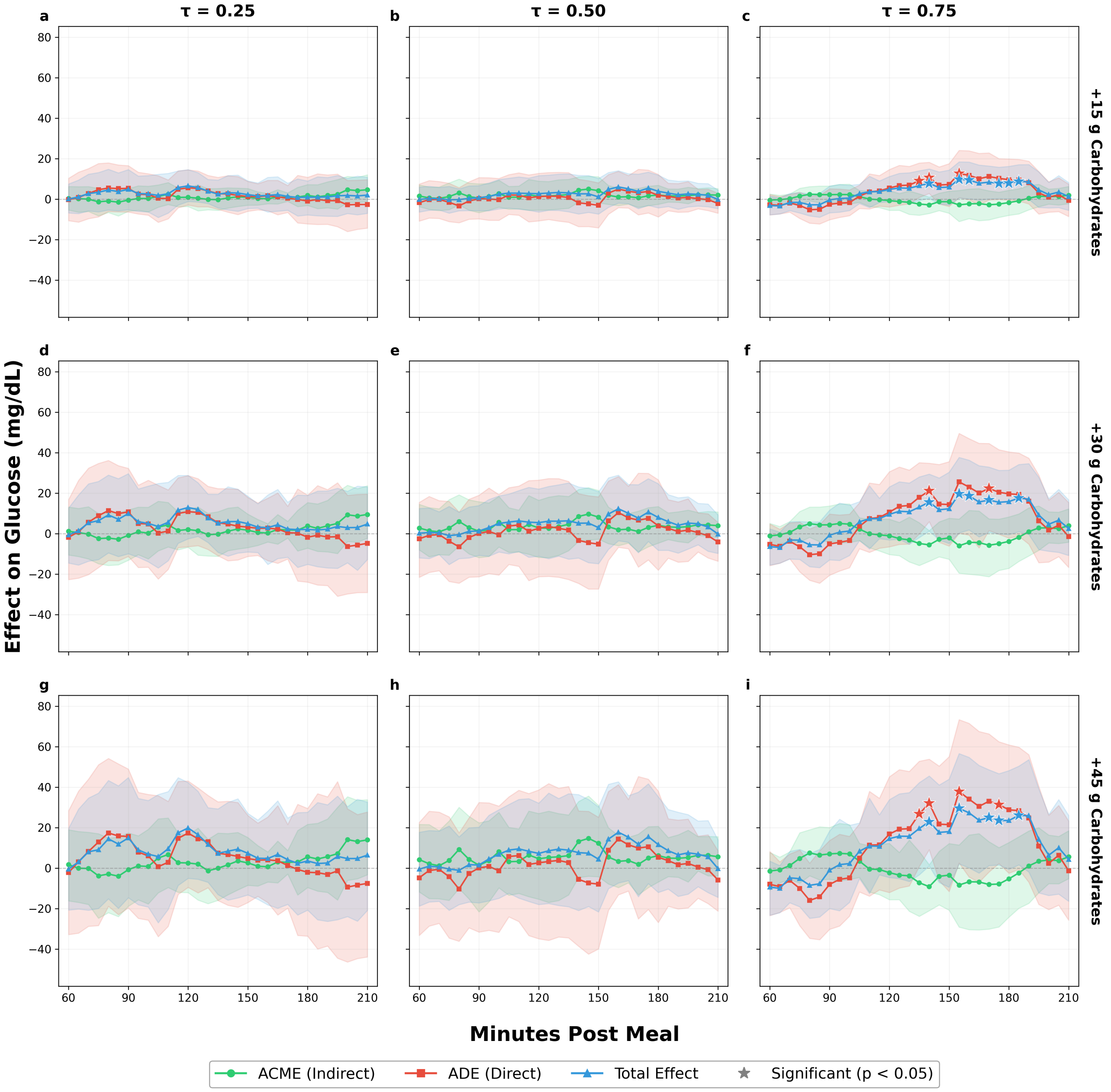


**Supplementary Figure 11: Quantile regression mediation effects — Lunch.** Rows: treatment contrasts (+15, +30, +45 g). Columns: $\tau \in \{0.25, 0.50, 0.75\}$.

##### D.4.2 Quantile Regression Effects

**Supplementary Table 10: QR causal mediation effects — Lunch ($N = 58$ meal observations, $+30$ g contrast).** ACME = Average Causal Mediation Effect; ADE = Average Direct Effect; Total = ACME + ADE. Est. = point estimate (mg/dL); CI = confidence interval. Significant results ($p < 0.05$) are shown in bold.

| | | ACME | | | ADE | | | Total | | |
|---|---|---|---|---|---|---|---|---|---|---|
| Time | Quantile | Est. | 95% CI | $p$ | Est. | 95% CI | $p$ | Est. | 95% CI | $p$ |
| 60 min | $\tau = 0.25$ | 1.14 | (-10.55, 13.18) | 0.83 | -1.78 | (-22.72, 16.85) | 0.90 | -0.64 | (-14.56, 12.68) | 0.94 |
| | $\tau = 0.50$ | 2.78 | (-8.03, 14.49) | 0.59 | -2.57 | (-21.60, 15.45) | 0.80 | 0.20 | (-11.97, 12.45) | 0.96 |
| | $\tau = 0.75$ | -1.14 | (-7.75, 5.07) | 0.75 | -5.27 | (-15.80, 4.79) | 0.32 | -6.42 | (-15.54, 2.37) | 0.16 |
| 90 min | $\tau = 0.25$ | -0.83 | (-13.15, 11.73) | 0.91 | 10.76 | (-11.25, 32.27) | 0.34 | 9.93 | (-9.94, 29.65) | 0.31 |
| | $\tau = 0.50$ | 1.17 | (-11.87, 15.76) | 0.89 | 0.11 | (-21.31, 18.99) | 1.00 | 1.28 | (-10.72, 13.88) | 0.86 |
| | $\tau = 0.75$ | 4.35 | (-3.71, 13.67) | 0.28 | -5.09 | (-20.56, 9.69) | 0.52 | -0.73 | (-12.94, 11.64) | 0.93 |
| 120 min | $\tau = 0.25$ | 2.05 | (-6.75, 12.01) | 0.66 | 10.85 | (-6.44, 28.70) | 0.21 | 12.90 | (-2.56, 28.78) | 0.10 |
| | $\tau = 0.50$ | 2.80 | (-11.79, 16.48) | 0.75 | 2.64 | (-21.29, 26.47) | 0.84 | 5.43 | (-11.17, 22.04) | 0.54 |
| | $\tau = 0.75$ | -1.07 | (-10.32, 7.16) | 0.80 | 10.60 | (-6.51, 30.60) | 0.25 | 9.53 | (-3.11, 23.11) | 0.11 |
| 150 min | $\tau = 0.25$ | 1.70 | (-6.92, 9.64) | 0.62 | 3.22 | (-11.26, 17.86) | 0.66 | 4.92 | (-9.07, 18.51) | 0.49 |
| | $\tau = 0.50$ | 8.17 | (-2.70, 21.37) | 0.18 | -5.20 | (-27.37, 18.20) | 0.66 | 2.97 | (-14.87, 20.71) | 0.74 |
| | $\tau = 0.75$ | -2.07 | (-12.67, 7.62) | 0.69 | 14.37 | (-6.16, 35.57) | 0.16 | 12.30 | (-4.88, 30.39) | 0.16 |
| 180 min | $\tau = 0.25$ | 3.84 | (-7.17, 15.36) | 0.44 | -1.75 | (-23.97, 18.99) | 0.92 | 2.09 | (-15.18, 18.90) | 0.79 |
| | $\tau = 0.50$ | 4.11 | (-4.68, 13.38) | 0.35 | 3.86 | (-18.05, 26.62) | 0.74 | 7.96 | (-8.26, 24.69) | 0.33 |
| | $\tau = 0.75$ | -3.83 | (-17.13, 8.26) | 0.57 | 19.66 | (-2.21, 40.41) | 0.07 | 15.83 | (-0.51, 31.37) | 0.05 |
| 210 min | $\tau = 0.25$ | 9.50 | (-2.28, 23.67) | 0.13 | -4.77 | (-29.09, 19.54) | 0.69 | 4.73 | (-12.79, 23.13) | 0.62 |
| | $\tau = 0.50$ | 3.95 | (-2.02, 10.70) | 0.24 | -4.15 | (-13.64, 5.51) | 0.43 | -0.20 | (-9.77, 9.22) | 0.99 |
| | $\tau = 0.75$ | 3.96 | (-4.21, 12.22) | 0.33 | -1.42 | (-16.72, 15.36) | 0.87 | 2.54 | (-10.79, 16.67) | 0.70 |

#### D.5 Snack

##### D.5.1 Mean Effects (LMER Outcome)

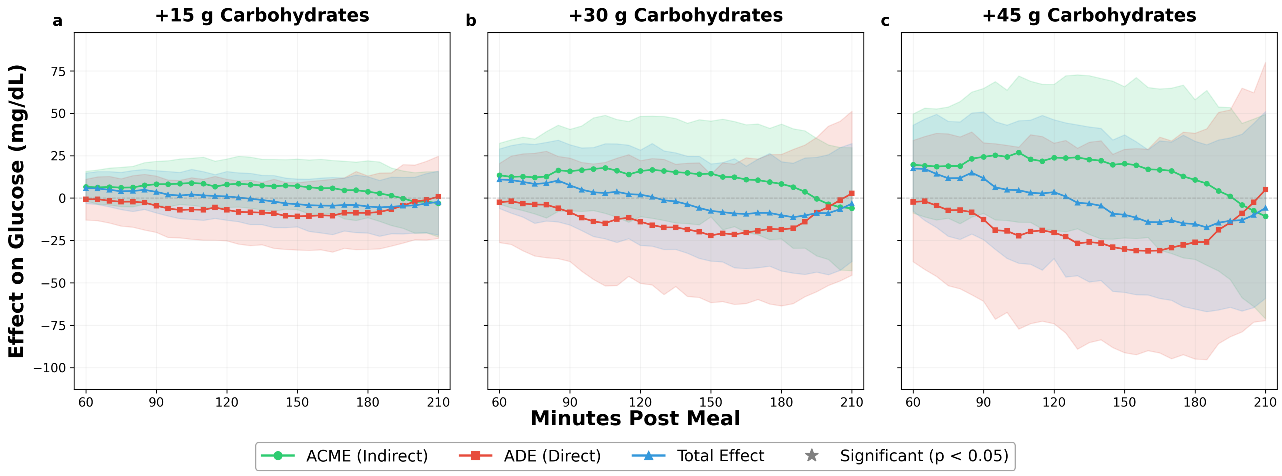


**Supplementary Figure 12: Mean causal mediation effects — Snack.** Panels show treatment contrasts ($+15$, $+30$, $+45$ g).

**Supplementary Table 11: LMER causal mediation effects — Snack ($N = 39$ meal observations, all contrast sizes).** ACME = Average Causal Mediation Effect; ADE = Average Direct Effect; Total = ACME + ADE. Est. = point estimate (mg/dL); CI = confidence interval. Significant results ($p < 0.05$) are shown in bold.

| | | ACME | | | ADE | | | Total | | |
|---|---|---|---|---|---|---|---|---|---|---|
| Time | Dose | Est. | 95% CI | $p$ | Est. | 95% CI | $p$ | Est. | 95% CI | $p$ |
| 60 min | $+15\,g$ | 6.58 | (-1.69, 15.81) | 0.13 | -0.77 | (-12.92, 11.12) | 0.93 | 5.81 | (-3.18, 14.75) | 0.21 |
| | $+30\,g$ | 13.44 | (-3.58, 32.36) | 0.14 | -2.48 | (-26.25, 20.45) | 0.82 | 10.96 | (-6.13, 29.00) | 0.20 |
| | $+45\,g$ | 19.70 | (-8.82, 49.60) | 0.17 | -2.28 | (-37.50, 34.02) | 0.88 | 17.42 | (-7.65, 42.99) | 0.19 |
| 90 min | $+15\,g$ | 8.15 | (-4.17, 21.67) | 0.18 | -4.51 | (-20.22, 13.01) | 0.58 | 3.64 | (-8.76, 16.18) | 0.54 |
| | $+30\,g$ | 15.85 | (-7.40, 40.58) | 0.19 | -8.33 | (-37.39, 23.60) | 0.62 | 7.52 | (-15.89, 31.48) | 0.56 |
| | $+45\,g$ | 24.31 | (-9.06, 64.70) | 0.17 | -12.61 | (-60.85, 32.61) | 0.63 | 11.70 | (-24.55, 50.85) | 0.57 |
| 120 min | $+15\,g$ | 7.97 | (-5.50, 23.07) | 0.26 | -7.01 | (-25.22, 11.12) | 0.45 | 0.97 | (-13.05, 15.69) | 0.91 |
| | $+30\,g$ | 15.94 | (-10.94, 48.28) | 0.26 | -13.94 | (-50.31, 22.09) | 0.45 | 2.00 | (-27.93, 28.32) | 0.88 |
| | $+45\,g$ | 23.86 | (-19.01, 67.07) | 0.25 | -20.35 | (-74.09, 32.42) | 0.47 | 3.51 | (-35.69, 46.22) | 0.89 |
| 150 min | $+15\,g$ | 7.15 | (-6.82, 23.05) | 0.35 | -10.75 | (-30.60, 9.11) | 0.29 | -3.60 | (-16.97, 11.25) | 0.63 |
| | $+30\,g$ | 14.47 | (-11.03, 45.44) | 0.31 | -22.05 | (-61.67, 14.00) | 0.30 | -7.58 | (-37.19, 20.16) | 0.60 |
| | $+45\,g$ | 20.30 | (-23.49, 65.73) | 0.36 | -30.14 | (-91.20, 28.79) | 0.32 | -9.84 | (-55.60, 37.31) | 0.67 |
| 180 min | $+15\,g$ | 3.81 | (-11.58, 22.01) | 0.68 | -8.77 | (-30.23, 13.07) | 0.43 | -4.96 | (-20.59, 12.61) | 0.56 |
| | $+30\,g$ | 8.36 | (-24.09, 43.66) | 0.63 | -18.51 | (-63.59, 25.78) | 0.41 | -10.15 | (-43.04, 21.46) | 0.54 |
| | $+45\,g$ | 10.73 | (-36.14, 58.07) | 0.67 | -26.03 | (-94.92, 38.26) | 0.45 | -15.29 | (-65.47, 33.54) | 0.53 |
| 210 min | $+15\,g$ | -3.08 | (-22.90, 15.58) | 0.75 | 0.83 | (-23.85, 24.70) | 0.92 | -2.25 | (-21.77, 15.94) | 0.82 |
| | $+30\,g$ | -6.01 | (-42.85, 29.76) | 0.73 | 2.73 | (-45.46, 51.26) | 0.93 | -3.29 | (-37.32, 32.20) | 0.85 |
| | $+45\,g$ | -10.79 | (-71.18, 49.42) | 0.70 | 4.96 | (-72.26, 80.10) | 0.93 | -5.83 | (-59.13, 51.08) | 0.81 |

##### D.5.2 Quantile Regression Effects

**Supplementary Table 12: QR causal mediation effects — Snack ($N = 39$ meal observations, $+30$ g contrast).** ACME = Average Causal Mediation Effect; ADE = Average Direct Effect; Total = ACME + ADE. Est. = point estimate (mg/dL); CI = confidence interval. Significant results ($p < 0.05$) are shown in bold.

| | | ACME | | | ADE | | | Total | | |
|---|---|---|---|---|---|---|---|---|---|---|
| Time | Quantile | Est. | 95% CI | $p$ | Est. | 95% CI | $p$ | Est. | 95% CI | $p$ |
| 60 min | $\tau = 0.25$ | -1.80 | (-82.04, 78.29) | 0.91 | 25.54 | (-12.44, 63.51) | 0.19 | 23.74 | (-43.39, 93.37) | 0.48 |
| | $\tau = 0.50$ | 6.40 | (-8.94, 23.98) | 0.42 | 6.68 | (-27.32, 38.71) | 0.70 | 13.08 | (-9.07, 34.10) | 0.23 |
| | $\tau = 0.75$ | **24.45** | **(10.30, 40.52)** | **<0.001** | **-19.90** | **(-39.10, -2.08)** | **0.03** | 4.54 | (-10.09, 18.14) | 0.57 |
| 90 min | $\tau = 0.25$ | 31.66 | (-11.65, 83.98) | 0.16 | -11.60 | (-75.14, 53.94) | 0.66 | 20.06 | (-48.00, 92.75) | 0.58 |
| | $\tau = 0.50$ | -0.17 | (-75.57, 72.44) | 0.98 | 21.76 | (-20.90, 65.07) | 0.34 | 21.60 | (-37.42, 83.24) | 0.51 |
| | $\tau = 0.75$ | 7.99 | (-53.43, 75.30) | 0.81 | -12.86 | (-51.67, 25.49) | 0.51 | -4.87 | (-48.90, 42.44) | 0.78 |
| 120 min | $\tau = 0.25$ | 12.78 | (-34.74, 64.92) | 0.59 | -5.04 | (-85.90, 73.94) | 0.93 | 7.74 | (-50.67, 62.22) | 0.76 |
| | $\tau = 0.50$ | 0.55 | (-84.62, 83.35) | 0.99 | 10.05 | (-45.87, 64.53) | 0.68 | 10.60 | (-56.06, 74.45) | 0.73 |
| | $\tau = 0.75$ | **21.43** | **(7.62, 37.24)** | **<0.001** | **-40.05** | **(-73.12, -8.59)** | **0.01** | -18.62 | (-46.97, 7.88) | 0.20 |
| 150 min | $\tau = 0.25$ | 9.08 | (-47.75, 65.44) | 0.74 | -7.25 | (-43.29, 28.85) | 0.67 | 1.83 | (-61.72, 61.89) | 0.92 |
| | $\tau = 0.50$ | 5.43 | (-80.81, 94.24) | 0.91 | -12.60 | (-62.73, 34.89) | 0.59 | -7.17 | (-55.20, 42.88) | 0.76 |
| | $\tau = 0.75$ | -1.03 | (-69.71, 65.83) | 0.98 | -26.28 | (-88.57, 38.94) | 0.41 | -27.31 | (-79.00, 25.94) | 0.34 |
| 180 min | $\tau = 0.25$ | 4.15 | (-105.44, 111.89) | 0.95 | 2.88 | (-62.34, 69.87) | 0.91 | 7.03 | (-54.13, 71.68) | 0.83 |
| | $\tau = 0.50$ | -2.84 | (-104.98, 98.58) | 0.96 | -7.92 | (-71.50, 57.55) | 0.79 | -10.76 | (-86.54, 63.57) | 0.75 |
| | $\tau = 0.75$ | -5.66 | (-129.65, 115.66) | 0.92 | -22.38 | (-116.17, 67.78) | 0.60 | -28.04 | (-100.00, 40.02) | 0.41 |
| 210 min | $\tau = 0.25$ | -8.56 | (-144.62, 115.94) | 0.93 | 22.15 | (-44.68, 88.18) | 0.49 | 13.58 | (-79.00, 102.83) | 0.77 |
| | $\tau = 0.50$ | -15.34 | (-125.28, 98.97) | 0.74 | 10.88 | (-45.75, 70.51) | 0.72 | -4.46 | (-79.94, 68.50) | 0.91 |
| | $\tau = 0.75$ | -15.71 | (-150.13, 123.53) | 0.80 | 4.97 | (-100.88, 104.98) | 0.92 | -10.74 | (-105.89, 81.27) | 0.83 |

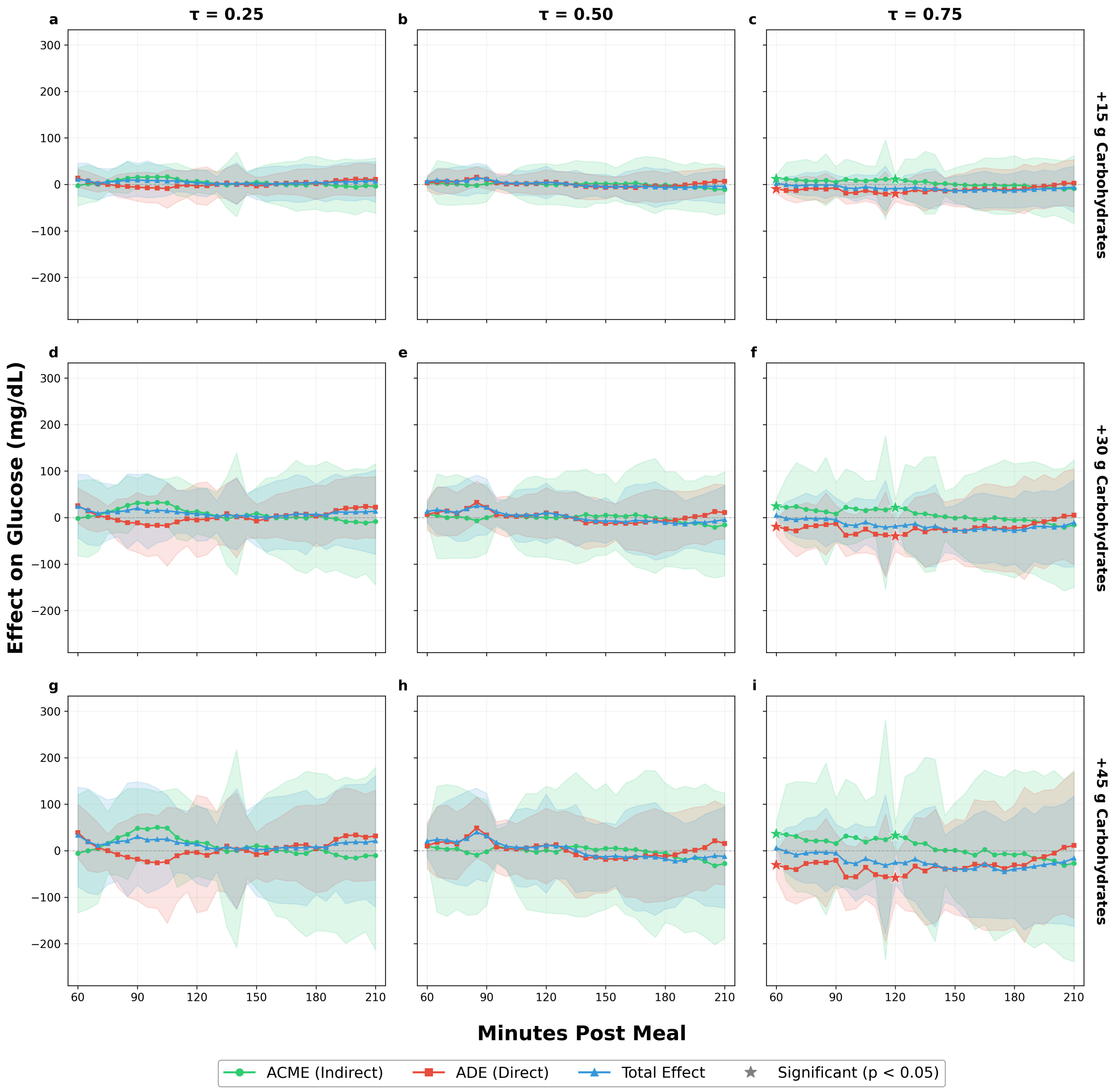


**Supplementary Figure 13: Quantile regression mediation effects — Snack.** Rows: treatment contrasts (+15, +30, +45 g). Columns: $\tau \in \{0.25, 0.50, 0.75\}$.

### E Quantile Regression Sensitivity Analysis

The mean-level (LMER) mediation results presented in the main text characterize the average causal mediation effect across the conditional glucose response distribution. To assess whether this average masks clinically relevant heterogeneity at the distributional tails, we re-estimated the mediation models using quantile regression (QR) for the outcome specification at $\tau \in \{0.25, 0.50, 0.75\}$, retaining the Tobit mediator model and npCBPS weights. Full QR figures and tables for each meal type appear in the Causal Mediation Analysis Results section above; here we consolidate the main findings across meal types.

Three patterns emerge from the distributional analysis. First, the $\tau = 0.75$ quantile consistently identifies the strongest and most clinically relevant effects. At the pooled level, the ADE reaches 19.73 mg/dL at 120 minutes ($p = 0.002$) and the total effect is 13.95 mg/dL ($p = 0.006$), nearly three times the non-significant LMER total effect of 5.17 mg/dL ($p = 0.13$) at the same time-point. By contrast, $\tau = 0.25$ effects are attenuated and generally non-significant across all meal types, confirming that clinically meaningful heterogeneity concentrates in the upper tail of the conditional glucose response distribution.

Second, the mediation structure at dinner is remarkably consistent across the distribution: incomplete insulin compensation (ADE exceeding ACME in magnitude) is present at every quantile examined, with the total effect growing monotonically from 10.79 mg/dL at $\tau = 0.25$ through 19.95 mg/dL at $\tau = 0.50$ to 22.03 mg/dL ($p = 0.04$) at $\tau = 0.75$. This consistency suggests that dinner bolusing is systematically miscalibrated across a broad range of the glucose response distribution, rather than being confined to a small subgroup of high-risk individuals.

Third, breakfast presents the sharpest contrast with dinner. The near-cancellation of direct and mediated effects persists across all three quantiles. At $\tau = 0.75$, both the ACME ($-24.58$ mg/dL, $p < 0.001$) and ADE (23.58 mg/dL, $p = 0.09$) are large, but their near-equality yields a total effect of $-1.00$ mg/dL ($p = 0.96$). This concentration of the strongest individual-pathway effects at the upper quantile suggests that the insulin compensation mechanism is most actively engaged among individuals who would otherwise experience the largest postprandial excursions, consistent with reactive bolusing behavior scaled to anticipated glycemic impact. Neither lunch nor snack produces significant effects at any quantile examined.

For automated insulin delivery (AID) systems, these findings suggest that meal-type-specific and quantile-aware dosing algorithms could improve glycemic outcomes beyond fixed carbohydrate-to-insulin ratios. In particular, dinner bolus calculations may benefit from upward adjustment to address the persistent ADE-ACME imbalance, especially for individuals identified as high glucose responders.

### F Model Selection and Ablation Results

**Supplementary Table 13: Architecture $\times$ optimizer comparison (averaged over penalties and seeds).** Values shown as mean $\pm$ SD. Sorted by balance score, then $R^2$.

| Architecture | Optimizer | Outcome $R^2$ | Balance Score |
|---|---|---|---|
| LSTM | adam | 0.343 ± 0.041 | 0.805 ± 0.179 |
| LSTM | adamw | 0.341 ± 0.041 | 0.801 ± 0.179 |
| CNN | adam | 0.361 ± 0.043 | 0.791 ± 0.145 |
| CNN | rmsprop | 0.382 ± 0.057 | 0.790 ± 0.143 |
| LSTM | rmsprop | 0.343 ± 0.035 | 0.778 ± 0.141 |
| CNN | adamw | 0.357 ± 0.040 | 0.765 ± 0.140 |

**Supplementary Table 14: Marginal contribution of each penalty layer.** $\Delta$ = mean metric with penalty enabled $-$ mean metric with penalty disabled, averaged across all other penalty combinations and seeds. Positive $\Delta R^2$ and $\Delta$ Balance are desirable.

| Penalty Layer | $\Delta R^2$ | $\Delta$ Balance | $\Delta$ Mediator $R^2$ |
|---|---|---|---|
| Linearization | -0.0129 | +0.0064 | -0.0512 |
| Balancing | -0.0424 | -0.2306 | -0.0688 |
| CI penalty | -0.0000 | +0.0037 | +0.0105 |
| Stability | -0.0054 | -0.0081 | +0.0008 |

**Supplementary Table 15: Top 10 penalization configurations from the ablation study (2 seeds each).** All $2^4 = 16$ combinations of the four penalty layers are evaluated. Architecture: CNN; optimizer: rmsprop. Values shown as mean ± SD across seeds. Training on combined 2018+2020 data. Treatment AUC ideal is 0.5 (no predictive power ⇒ balanced embedding).

| Rank | Configuration | Outcome $R^2$ | Balance Score | Mediator $R^2$ | Treatment AUC |
|---|---|---|---|---|---|
| 1 | Linearization + Stability | 0.356 ± 0.000 | 0.979 ± 0.000 | 0.428 ± 0.000 | 0.490 ± 0.000 |
| 2 | Linearization | 0.361 ± 0.000 | 0.964 ± 0.000 | 0.428 ± 0.000 | 0.518 ± 0.000 |
| 3 | Linearization + CI | 0.367 ± 0.000 | 0.963 ± 0.000 | 0.460 ± 0.000 | 0.482 ± 0.000 |
| 4 | CI + Stability | 0.382 ± 0.000 | 0.950 ± 0.000 | 0.495 ± 0.000 | 0.475 ± 0.000 |
| 5 | CI penalty | 0.379 ± 0.000 | 0.946 ± 0.000 | 0.501 ± 0.000 | 0.473 ± 0.000 |
| 6 | Baseline (none) | 0.370 ± 0.000 | 0.928 ± 0.000 | 0.506 ± 0.000 | 0.464 ± 0.000 |
| 7 | Stability | 0.373 ± 0.000 | 0.847 ± 0.000 | 0.510 ± 0.000 | 0.424 ± 0.000 |
| 8 | Balancing + CI + Stability | 0.331 ± 0.000 | 0.742 ± 0.000 | 0.433 ± 0.000 | 0.371 ± 0.000 |
| 9 | Balancing + Stability | 0.329 ± 0.000 | 0.734 ± 0.000 | 0.420 ± 0.000 | 0.367 ± 0.000 |
| 10 | Balancing + CI | 0.319 ± 0.000 | 0.732 ± 0.000 | 0.435 ± 0.000 | 0.366 ± 0.000 |

**Supplementary Table 16: Top 10 autoencoder configurations ranked by balance score then outcome $R^2$ (20 total configurations, 2 seeds each).** Each configuration is a combination of encoder architecture, optimizer, and penalty regime. Balance score $= 1 - |0.5 - \text{AUC}_{\text{treatment}}| \times 2$ (1.0 = ideal). Values shown as mean ± SD across seeds. Training on combined 2018+2020 data.

| Rank | Architecture | Optimizer | Penalty | Outcome $R^2$ | Balance Score |
|---|---|---|---|---|---|
| 1 | CNN | rmsprop | Linear only | 0.431 ± 0.005 | 0.972 ± 0.012 |
| 2 | CNN | adam | Linear only | 0.402 ± 0.005 | 0.965 ± 0.007 |
| 3 | LSTM | adam | None | 0.378 ± 0.004 | 0.960 ± 0.006 |
| 4 | LSTM | adamw | None | 0.376 ± 0.004 | 0.956 ± 0.006 |
| 5 | LSTM | rmsprop | None | 0.388 ± 0.004 | 0.955 ± 0.007 |
| 6 | CNN | adam | None | 0.417 ± 0.004 | 0.945 ± 0.007 |
| 7 | CNN | adamw | None | 0.407 ± 0.004 | 0.940 ± 0.006 |
| 8 | CNN | rmsprop | None | 0.463 ± 0.006 | 0.934 ± 0.009 |
| 9 | LSTM | rmsprop | Linear + Balance | 0.323 ± 0.004 | 0.725 ± 0.007 |
| 10 | CNN | adam | Linear + Balance | 0.338 ± 0.004 | 0.720 ± 0.006 |

### G External Validation: DiaTrend Cohort

This section reports the data characteristics and causal mediation results for the independent DiaTrend cohort (Prioleau et al., 2023) used to replicate the primary direct-effect finding (see main text, "Replication in an independent cohort," and main-text Figure 4). Meal windows, treatment, mediator, outcome, the CLAE embedding, npCBPS weighting, and the mediation estimators are defined identically to the OhioT1DM analysis. The analyzed cohort comprises 54 adults with type 1 diabetes; the causal mediation analysis was performed on the held-out test set of 1,890 demographics-complete meal events.

**Supplementary Table 17: DiaTrend data characteristics by cohort and meal type.** Summary statistics for carbohydrate intake (treatment), insulin bolus (mediator), and pre-meal glucose stratified by cohort and meal category. N = number of meal observations; SD = standard deviation; values are mean ± SD; % Zero Bolus = percentage of meals with no insulin bolus. The causal mediation analysis was performed on the held-out test set ($N = 1{,}890$ demographics-complete meal observations).

| | | | Carbs (g) | | Bolus (U) | | % Zero | Glucose (mg/dL) |
|---|---|---|---|---|---|---|---|---|
| Cohort | Meal Type | N | Mean | SD | Mean | SD | Bolus | Mean ± SD |
| 1 | Breakfast | 105 | 35.1 | 24.3 | 4.50 | 2.80 | 0.0 | 215.2 ± 59.6 |
| | Lunch | 551 | 30.5 | 17.4 | 4.50 | 3.10 | 0.0 | 170.4 ± 53.7 |
| | Dinner | 495 | 29.9 | 21.8 | 4.90 | 4.50 | 0.0 | 175.7 ± 57.1 |
| | Snack | 1332 | 32.2 | 21.2 | 6.10 | 5.70 | 0.2 | 183.1 ± 59.4 |
| 2 | Breakfast | 859 | 40.5 | 22.6 | 7.00 | 4.80 | 0.2 | 179.3 ± 69.1 |
| | Lunch | 1812 | 44.6 | 24.4 | 8.90 | 6.10 | 0.0 | 187.6 ± 69.3 |
| | Dinner | 2594 | 44.7 | 24.2 | 9.50 | 6.50 | 0.1 | 198.1 ± 70.5 |
| | Snack | 2296 | 40.8 | 25.5 | 8.40 | 5.90 | 0.2 | 211.4 ± 75.0 |

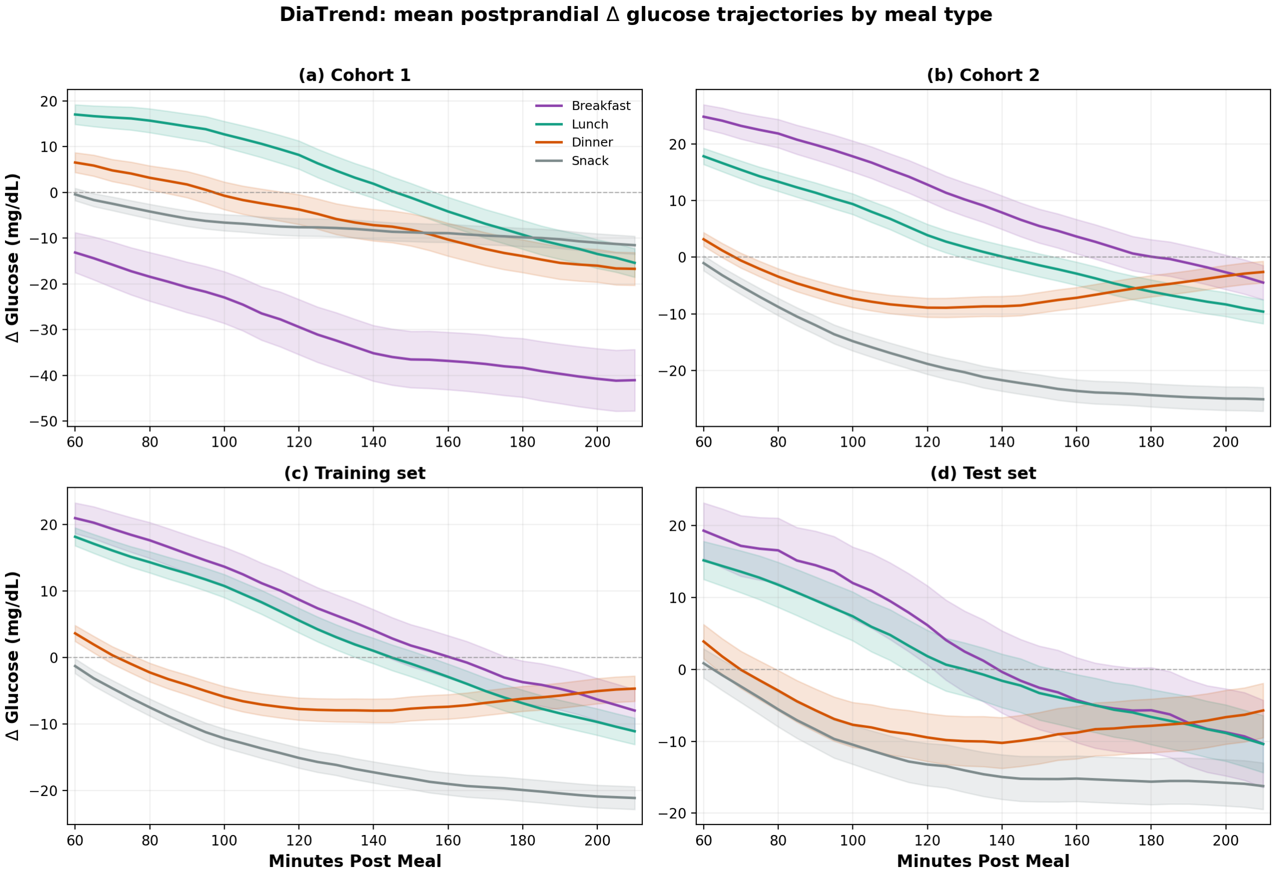


**Supplementary Figure 14: DiaTrend postprandial Δ glucose trajectories by meal type.** Mean ± 1 SE glucose excursion ($\Delta G$) from 60 to 210 minutes post-meal. (**a**, **b**) Trajectories stratified by device cohort; (**c**, **d**) trajectories stratified by training and test sets. Dinner and lunch show the largest sustained excursions, paralleling the OhioT1DM pattern.

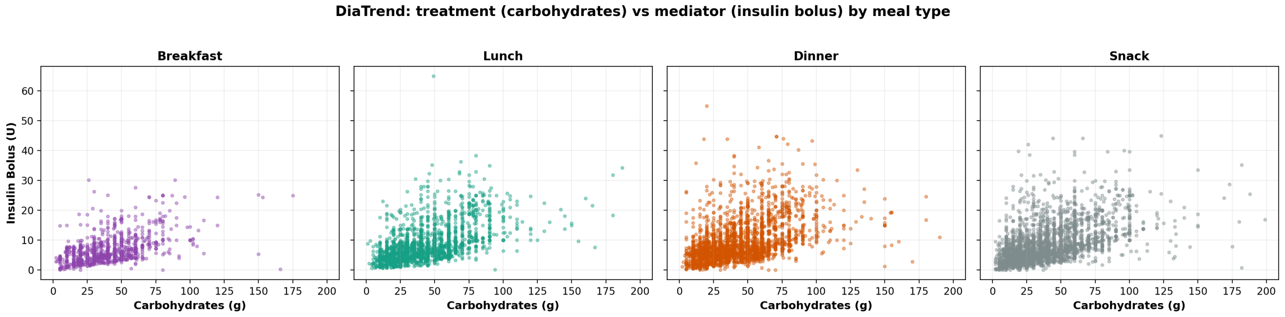


**Supplementary Figure 15: DiaTrend treatment-mediator relationship by meal type.** Scatter plots of carbohydrate intake (treatment) versus insulin bolus (mediator) for each meal type in the DiaTrend cohort, parallel to the OhioT1DM relationship in Supplementary Figure 1.

**Supplementary Table 18: DiaTrend — Pooled (all meals): LMER causal mediation effects across treatment doses** ($N = 1{,}890$ **meal observations).** ACME = average causal mediation effect (through insulin); ADE = average direct effect; Total = ACME + ADE. Est. in mg/dL; 95% CI and $p$ from quasi-Bayesian approximation. Significant ($p < 0.05$) in bold.

| | | ACME | | | ADE | | | Total | | |
|---|---|---|---|---|---|---|---|---|---|---|
| Time | Dose | Est. | 95% CI | $p$ | Est. | 95% CI | $p$ | Est. | 95% CI | $p$ |
| 60 min | +15 g | -0.11 | (-1.05, 0.77) | 0.80 | 0.59 | (-1.34, 2.57) | 0.55 | 0.48 | (-1.06, 2.14) | 0.59 |
| | +30 g | -0.19 | (-1.93, 1.55) | 0.85 | 1.14 | (-2.51, 4.72) | 0.53 | 0.95 | (-1.97, 4.13) | 0.55 |
| | +45 g | -0.26 | (-2.84, 2.41) | 0.83 | 1.81 | (-3.51, 7.51) | 0.53 | 1.55 | (-3.40, 6.58) | 0.53 |
| 90 min | +15 g | -0.64 | (-1.72, 0.47) | 0.22 | 1.13 | (-0.89, 3.32) | 0.32 | 0.49 | (-1.36, 2.59) | 0.60 |
| | +30 g | -1.30 | (-3.32, 0.88) | 0.22 | 2.33 | (-2.29, 6.73) | 0.28 | 1.03 | (-2.86, 4.89) | 0.58 |
| | +45 g | -1.88 | (-5.16, 1.35) | 0.25 | 3.61 | (-2.95, 10.23) | 0.27 | 1.73 | (-4.31, 7.26) | 0.55 |
| 120 min | +15 g | -0.92 | (-2.03, 0.17) | 0.09 | **2.70** | **(0.23, 5.16)** | **0.03** | 1.78 | (-0.22, 3.87) | 0.09 |
| | +30 g | -1.83 | (-4.27, 0.48) | 0.11 | **5.29** | **(0.63, 10.11)** | **0.02** | 3.46 | (-0.72, 7.59) | 0.13 |
| | +45 g | -2.65 | (-6.15, 0.78) | 0.14 | **7.87** | **(0.77, 15.24)** | **0.04** | 5.22 | (-0.94, 11.32) | 0.11 |
| 150 min | +15 g | -0.82 | (-2.06, 0.31) | 0.16 | **3.69** | **(1.08, 6.37)** | **0.006** | **2.88** | **(0.60, 5.27)** | **0.01** |
| | +30 g | -1.54 | (-3.95, 0.75) | 0.21 | **7.20** | **(1.70, 12.17)** | **0.006** | **5.66** | **(1.22, 9.89)** | **0.02** |
| | +45 g | -2.45 | (-5.92, 1.03) | 0.19 | **10.99** | **(3.38, 18.53)** | **0.004** | **8.54** | **(1.90, 15.39)** | **0.01** |
| 180 min | +15 g | -0.55 | (-1.73, 0.58) | 0.37 | **3.74** | **(0.95, 6.38)** | **0.004** | **3.19** | **(1.00, 5.52)** | **0.01** |
| | +30 g | -1.18 | (-3.40, 1.26) | 0.31 | **7.66** | **(2.57, 12.36)** | **0.002** | **6.48** | **(1.94, 10.81)** | **0.002** |
| | +45 g | -1.77 | (-5.39, 1.68) | 0.36 | **11.69** | **(4.38, 19.53)** | **0.002** | **9.92** | **(3.27, 16.53)** | **<0.001** |
| 210 min | +15 g | -0.16 | (-1.33, 1.05) | 0.78 | **4.35** | **(1.68, 6.93)** | **<0.001** | **4.19** | **(1.79, 6.53)** | **<0.001** |
| | +30 g | -0.31 | (-2.70, 2.03) | 0.77 | **8.53** | **(3.59, 14.06)** | **<0.001** | **8.22** | **(3.82, 12.72)** | **<0.001** |
| | +45 g | -0.56 | (-4.16, 2.82) | 0.80 | **12.85** | **(4.90, 20.78)** | **<0.001** | **12.29** | **(5.62, 18.70)** | **<0.001** |
